# Longitudinal analysis reveals elevation then sustained higher expression of autoantibodies for six months after SARS-CoV-2 infection

**DOI:** 10.1101/2022.05.04.22274681

**Authors:** Nahid Bhadelia, Alex Olson, Erika Smith, Katherine Riefler, Jacob Cabrejas, Maria-Jose Ayuso, Katherine Clarke, Rachel R. Yuen, Nina H. Lin, Zachary J. Manickas-Hill, Ian Rifkin, Andreea Bujor, Manish Sagar, Anna C. Belkina, Jennifer E. Snyder-Cappione

**Affiliations:** Department of Medicine, Boston University School of Medicine, Boston, MA, USA; Center for Emerging Infectious Diseases Policy & Research, Boston University School of Medicine, Boston, MA, USA; National Emerging Infectious Diseases Laboratories (NEIDL), Boston University, Boston, MA, USA; Department of Microbiology; Boston University School of Medicine, Boston, MA, USA; Ragon Institute of MGH, MIT, and Harvard, Boston, MA, USA; Renal Section, Department of Medicine, Boston University School of Medicine, Boston, MA, USA; Renal Section, Department of Medicine, VA Boston Healthcare System, Boston, MA, USA; Rheumatology, Arthritis and Autoimmune Diseases Center, Boston University Medical Center, Boston, MA, USA; Flow Cytometry Core Facility, Boston University School of Medicine, Boston, MA, USA

## Abstract

High autoantibody levels are found in individuals hospitalized for COVID-19. The temporal trajectories and levels of these autoantibodies months into convalescence after SARS-CoV-2 infection are unclear. It is also unknown if the composite autoantibody signatures of convalescent SARS-CoV-2-infected individuals resemble those with diagnosed autoimmune diseases. We measured the circulating levels of 17 autoantibodies associated with autoimmune connective tissue diseases from SARS-CoV-2 hospitalized and outpatient participants, as well as from individuals with scleroderma (SSc), systemic lupus erythematosus (SLE), and uninfected pre-pandemic controls. Seven of the 17 autoantibodies measured were higher in hospitalized and/or outpatient SARS-CoV-2 individuals an average of six months after symptom onset compared with controls, with multivariate analyses revealing links between SARS-CoV-2 infection and positivity of SSB-La, Sm, Proteinase 3, Myleoperoxidase, Jo-1, and Ku reactive IgG six months post-symptom onset. Autoantibody levels from SARS-CoV-2 infected individuals were followed over time from initial symptom onset for an average of six months, and different temporal autoantibody trajectories were classified. A ‘negative, then positive’ expression pattern was found for at least one autoantibody in 18% of the outpatient and 53% of the hospitalized participants, indicating initiation and durable expression of self-reactive immune responses post-infection, particularly with severe acute illness. Analysis of individual participant autoantibody expression patterns revealed similar patterns between pre-pandemic and convalescent SARS-CoV-2 infected groups that are distinct from participants with both the SSc and SLE. As autoantibody positivity can occur years prior to autoimmune disease onset, the possibility that SARS-CoV-2-associated autoantibodies are a herald of future autoimmune disorders requires further investigation.

**One Sentence Summary:** Autoantibody levels rise after acute SARS-CoV-2 infection and remain elevated for at least six months after symptom onset in participants with mild or severe COVID-19.

## Introduction

Individuals hospitalized with severe COVID-19 express autoantibodies reactive with a range of self-epitopes, including interferons and other cytokines, DNA-associated antigens, phospholipids, gastrointestinal antigens, and coagulants [1–4]. These autoantibodies can have functional impact [1, 3, 4] and correlations are reported between autoantibody levels and COVID-19 severity [1, 5]. The proportions of autoantibodies that are newly induced after SARS-CoV-2 infection is unclear to date and the durability of autoantibody expression months after resolution of acute illness is currently unknown.

New onset inflammatory diseases with putative autoimmune or inflammatory drivers have been diagnosed after SARS-CoV-2 infection, such as pediatric inflammatory multisystemic syndrome (PIMS) or multisystem inflammatory syndrome in children (MIS-C), as well as the autoimmune conditions such as Guillain-Barre syndrome, Immune Thrombocytopenic Purpura, and autoimmune hemolytic anemia [6–11]. Pediatric Type I and Type 2 diabetes cases are rising since the start of the pandemic [12, 13]; also, unusually high cases of new onset diabetes in children and adults are reported months after infection with SARS-CoV-2 [14, 15]. These new disease diagnoses contribute to the growing literature illuminating the frequencies and morbidities of individuals afflicted with post-acute sequalae of SARS-CoV-2 (PASC) [16]. A recent cohort also reported correlation between autoantibodies and the presence of PASC symptoms [17].

To gain new insight into the links between autoimmunity and SARS-CoV-2 infection, we measured 17 autoantibodies found in connective tissue autoimmune diseases and related conditions, such as Sjogren’s syndrome, scleroderma, systemic lupus erythematosus, and Raynaud’s phenomenon, from SARS-CoV-2 infected individuals with either mild or severe initial illness (outpatient and hospitalized). We cross-sectionally compared autoantibody levels from both early and late time points of the outpatient and hospitalized SARS-CoV-2 infected participants (≤30 or ≥90 days post symptom onset) with pre-pandemic (uninfected) controls and two autoimmune participant groups, scleroderma (SSc) and systemic lupus erythematosus (SLE). We performed multivariate analyses to determine how previous SARS-CoV-2 infection associated with autoantibody positivity as compared to the demographics and co-morbidities of the participants. Also, we determined different patterns of changes in individual autoantibody levels in all SARS-CoV-2 infected individuals over time (two-five time points per participant, spanning an average of six months) to examine the longitudinal trajectory of autoantibodies months after symptom onset. In addition, we performed partial least squares discriminant analysis (PLS-DA) to compare the individual autoantibody expression signatures of SARS-CoV-2 infected individuals months into convalescence with both uninfected controls and individuals with established autoimmune diseases.

## Materials and Methods

### Study populations

Participant samples included plasma (pre-pandemic, SARS-CoV-2 infected, SSc) or serum (SLE). Samples from the SARS-CoV-2 infected groups were obtained from two COVID-19 biorepositories. 31 samples of PCR confirmed inpatients admitted between March to November 2020 were obtained from Boston Medical Center’s (BMC) discarded clinical samples biorepository [18]. The remaining SARS-CoV-2 samples were obtained from the Mass General Brigham (MGB) prospective clinical biorepository for patients recruited either in the outpatient clinic or inpatient wards between March and June 2020. Inclusion criteria for samples from both these sources included those who had at least two longitudinal samples spanning from acute (within 10 days of symptom onset) to convalescent (more than 30 days after symptom onset) phase of illness. Cases in all analysis identified as “inpatient/hospitalized” or “outpatient” refers to whether or not they required hospitalization during acute illness but samples from these individuals were drawn both during acute illness and after recovery. Pre-pandemic samples were from other existing studies and were biobanked prior to December 2019. The samples of participants with Scleroderma were obtained from an existing database of SSc patients from Boston University School of Medicine (SCAR Clinical Trial database) and included individuals with varying stages and types of disease (early and late SSc, limited and diffuse subtypes, with and without interstitial lung disease or pulmonary hypertension). All scleroderma patients fulfilled the new ACR/EULAR 2013 classification criteria and were classified as limited or diffuse SSc according to LeRoy classification [19]. Systemic lupus erythematosus samples were previously biobanked and were obtained under the following protocols: Autoimmune Kidney Research Studies and Patient Registry and the Autoimmune Kidney Research Repository approved by the Boston University Medical Center Institutional Review Board. All samples used in these studies were from lupus patients that fulfilled at least four of the 11 American College of Rheumatology revised criteria for the classification of SLE. Inclusion criteria for participants with lupus included the following: (1) participants must be >18 years of age, (2) participants with a diagnosis of autoimmune disease with or without kidney disease, and (3) participants must be able to read and understand the consent form. Exclusion criteria for lupus participants included the following: (1) Cognitive impairment who are unable to consent, (2) Diagnosis of any malignancy within the previous 2 years, excluding adequately treated squamous cell skin cancer, basal cell carcinoma, and carcinoma in situ, and (3) Presence of other co-morbid illnesses with an estimated median life expectancy < 5 years.

### Ethics

Human subjects review and permissions were obtained from the Boston University School of Medicine’s Institutional Review Board for all cohorts. Samples and minimal clinical data were deidentified by biorepositories at collection (MGB biorepository) or at sharing (pre-pandemic, SSc and SLE participants). Data from BMC discarded biorepository participants were extracted from medical records and then deidentified for study analyses.

### Autoantibody Measurements

The MILLIPLEX MAP Human Autoimmune Autoantibody Panel (Cat. No. HAIAB-10K, MilliporeSigma) was used to determine all autoantibody levels. Kit readouts were measured using the Magpix (Luminex) instrument containing xPONENT 4.2 software (Boston University Medical Center Analytical Core Facility). Antibody levels are expressed as Mean Fluorescence Intensity (MFI) and positive autoantibody hits in individuals were determined. Specifically, to determine the presence (hit) or absence of an autoantibody, first, the mean fluorescence intensity (MFI) from all samples measured in this study were cross-sectionally compared, including all pre-pandemic, SARS-CoV-2, SSc, and SLE individuals, with the latter two participant groups serving as positive control to guide cutoff decisions. Second, the MFIs in one participant over multiple time points (SARS-CoV-2 infected only) were analyzed, as for most autoantibody readouts, the MFI values were similar between early and later time points (either always positive, or always negative) and therefore gave an often-clear indication of positive versus negative results. One MFI cutoff value was determined per autoantibody and this same threshold was applied to all participant groups and samples (all listed in Supplemental Figure 2). Autoantibody hit thresholds were determined in this manner to aid detection of MFI shifts due to molecular mimicry.

### Statistical Analyses

Univariate analyses were conducted between groups comparing the number of autoantibody hits and log transformed MFIs using non-parametric Mann-Whitney U-test (Two stage step-up, false discovery rate 5%) or Kruskal-Wallis test (with Dunn correction for multiple comparisons). Poisson or least squares multiple linear regression was performed if the univariate comparisons between groups were statistically different (p-value <0.05). Multiple linear regression was conducted with the number of total autoantibody hits (Poisson) or differences in log mean fluorescence intensities (least squares) among each autoantibody as the dependent variables. Available demographics and comorbidities (age, sex, race, ethnicity, diabetes, heart disease, hypertension, lung disease, autoimmune disease, HIV status, and immunosuppressed) were included and removed in a backwards stepwise manner to establish the most parsimonious model to explain the data. All reported data meets the assumptions of the model unless otherwise stated. Partial Least Squares discriminant analysis (PLSDA) was conducted using PLS Toolbox (Eigenvector Research, Inc.) package for MATLAB (MathWorks) as previously described [20] to test whether a set of autoantibodies can distinguish between four participant groups (pre-pandemic, convalescent SARS-CoV-2 infected, SSc, and SLE). Briefly, raw fluorescence data was normalized by Z-score before application of the algorithm. The importance of each parameter to the overall model prediction was quantified using variable importance in projection (VIP) score. A PLSDA model was built based on 17 autoantibody measurements in the plasma. Cross-validation was performed with one-third of the dataset and VIP scores were calculated. The number of latent variables (LVs) was chosen to minimize cumulative error over all predictions. A VIP score >1 (above average contribution) was considered important for model performance and prediction. Model confidence was calculated by randomly permuting Y 100 times and rebuilding the model to form a distribution of error for these randomly generated models, then comparing the model to this distribution with the Mann–Whitney U-test to determine the significance of the model. Other statistical analyses were carried out using GraphPad version 9 or Matlab. All p-values are 2-sided unless indicated.

## Results

### Participant Characteristics

To examine possible connections between SARS-CoV-2 infection and autoantibodies, plasma or serum was analyzed from five groups (**Table 1**): (1) SARS-CoV-2 unexposed (collected ‘pre-pandemic’, before December 2019), (2) inpatient (hospitalized) SARS-COV-2 infected, (3) outpatient SARS-COV-2 infected, (4) scleroderma (SSc), and (5) systemic lupus erythematosus (SLE). Specimens from SARS-CoV-2 unexposed (pre-pandemic) individuals (n=52) had an average age of 52.3 years, and 37 (71%) were male. Inpatient (hospitalized) SARS-CoV-2 infected individuals were all afflicted with moderate or severe COVID-19 (n=40) with an average age of 58.7 years, and 26 (65%) were male. Multiple inpatient samples were collected per participant over time (2-4) ranging from 0-254 days after onset of symptoms. Twenty-two (55%) of the individuals in this cohort required intensive care unit level care during their admission, one was HIV positive, and no patients died. Outpatient SARS-CoV-2 infected individuals (n=33) were all symptomatic and an average age of 43, and nine (27.2%) were male, and none were hospitalized during the acute course of their infection. Multiple SARS-CoV-2 infected outpatient samples were collected per person over time (2-5) ranging from 1-260 days after onset of symptoms. There are demographic differences between the hospitalized COVID-19 patients who were predominantly male (65%) and the SARS-CoV-2 infected outpatient individuals who were predominantly white (96.9%). Outpatient SARS-CoV-2 infected individuals had fewer comorbidities compared to those requiring hospital admission. Scleroderma (SSc) individuals (n=40) included 20 with limited and 20 with diffuse disease, with an average age of 54.3 years, and 11 (27.5%) were male. The systemic lupus erythematosus (SLE) group (n=19) had an average age of 41.2 years, and five (26%) were male. No individuals from the SSc or SLE groups were taking immunosuppressants at the time of sample collection.

**Table 1:** Subject Demographic and Illness Features.

|  | **Prepandemic** | **Inpatient**<br>**SARS-CoV2** | **Outpatient**<br>**SARS-CoV2** | **Systemic**<br>**Lupus**<br>**Erythematous** | **Scleroderma** |
| --- | --- | --- | --- | --- | --- |
| **Total (N/%)=** | n=52 (%) | n=40 (%) | n=33 (%) | n=19 (%) | n=40 (%) |
| **Age in years (mean)** | 52.3 | 58.7 | 43 | 41.2 | 54.3 |
| **Sex (male)** | 37 (71%) | 26 (65%) | 9 (27.2%) | 5 (26%) | 11 (27.5%) |
| **Race** |  |  |  |  |  |
| Black | 23 (44.2%) | 22 (55%) | 0 | 3 (15.8%) | 1 (2.5%) |
| Asian | 0 | 3 (7.5%) | 0 | 2 (10.5%) | 1 (2.5%) |
| White | 27 (51.9%) | 6 (15%) | 32 (96.9%) | 8 (42%) | 38 (95%) |
| Other or not available | 2 (3.8%) | 9 (22.5%) | 1 (3%) | 6 (31.6%) | 0 |
| Ethnicity (Hispanic) | 5 (9.6%) | 12 (30%) | 1 (3%) | 6 (31.5%) | 0 |
| **Illness features** | N/A |  |  | N/A | N/A |
| Symptomatic |  | 40 (100%) | 33 (100%) |  |  |
| **Average follow up time**<br>**after symptom onset**<br>**(days)** |  | 107 | 199 |  |  |
| Last sample <=30 days |  | 1 (2.5%) | 0 |  |  |
| >30 and <=90 days |  | 19 (47.5%) | 1 (3%) |  |  |
| >90 and <=180 days |  | 13 (32.5%) | 6 (18%) |  |  |
| > 180 days |  | 7 (17.5%) | 26 (79%) |  |  |
| **Length of hospitalization** |  |  |  |  |  |
| Mean |  | 32.1 days | N/A |  |  |
| Median |  | 31.5 days | N/A |  |  |
| **ICU status (YES)** | N/A | 22 (55%) | N/A | N/A | N/A |
| **Medical Comorbidities** |  |  |  | N/A | N/A |
| Diabetes | 8 (15.4%) | 19 (43%) | 1 (3%) |  |  |
| Heart disease | 5 (9.6%) | 17 (39%) | 3 (9.1%) |  |  |
| Lung disease | 9 (17.3%) | 19 (43%) | 5 (15.1%) |  |  |
| Autoimmune disease | 15 (28.8%) | 3 (7%) | 3 (9.1%) |  |  |
| Immunosuppressed | 3 (5.8%) | 9 (20%) | 2 (6%) |  |  |
| HIV (well controlled) | 8 (15.4%) | 1 (2.5%) | 1 (3%) |  |  |
| Hypertension | 14 (26.9%) | 30 (68%) | 2 (6%) |  |  |
| Obesity (BMI>30) | N/A | 14 (3.2%) | 8 (24%) |  |  |
| ESRD | N/A | 7 (16%) | 0 |  |  |

### Seven of the 17 autoantibodies measured are higher in SARS-CoV-2 infected individuals (inpatient and/or outpatient) an average of six months post symptom onset compared with pre-pandemic controls

We first compared the autoantibody levels (MFI) between samples from pre-pandemic controls and SARS-CoV-2-infected individuals parsed into four groups by hospitalization status (inpatient and outpatient) and time post-symptom onset (≤30 days (acute) or ≥90 days (convalescent)). In agreement with others [1–5] the average MFIs of many autoantibodies (10 of the 17 measured) were significantly higher in participants who were acutely hospitalized for COVID-19 compared with pre-pandemic controls; in contrast, the acute outpatient samples were significantly higher for only three autoantibodies compared with pre-pandemic controls, SSB/La, Proteinase 3, and Jo-1 (**Figure 1A and Supplemental Figure 1A**). Strikingly, of the SARS-CoV-2 convalescent samples, the inpatient (average 168 days post symptom onset (PSO)) and/or the outpatient (average 200 days PSO) groups possessed higher levels of seven autoantibodies compared with controls: SSB/La, RNP/Sm, Proteinase 3, β-2-Glycoprotein, Scl-70, Ku, and PL-12 (**Figure 1A**). Also, compared with pre-pandemic controls: (1) SSB/La was higher among all four SARS-CoV-2 infected sample groups; (2) β-2-Glycoprotein was higher among convalescent inpatient and not outpatient individuals; and (3) RNP/Sm, Proteinase 3, Scl-70, and Ku were all higher in the outpatient convalescent but not inpatient samples as compared with pre-pandemic controls (**Figure 1A**). Three autoantibodies (Myeloperoxidase, CENP-A, and Jo-1) were significantly higher in acute SARS-CoV-2 but not convalescent samples as compared with controls (**Figure 1A**). Six autoantibodies (SSA-RO60, SSA-RO52, Sm, PCNA, CENP-B, and PM-Scl-100) were similar in average mean fluorescence intensity between all SARS-CoV-2 sample groups and controls (**Supplemental Figure 1A**). The 17 autoantibodies were also measured from individuals with diagnosed autoimmune diseases, either scleroderma (SSc) and systemic lupus erythematosus (SLE), and those levels are shown (**Figure 1A** and **Supplemental Figure 1A**).

**Figure 1.**
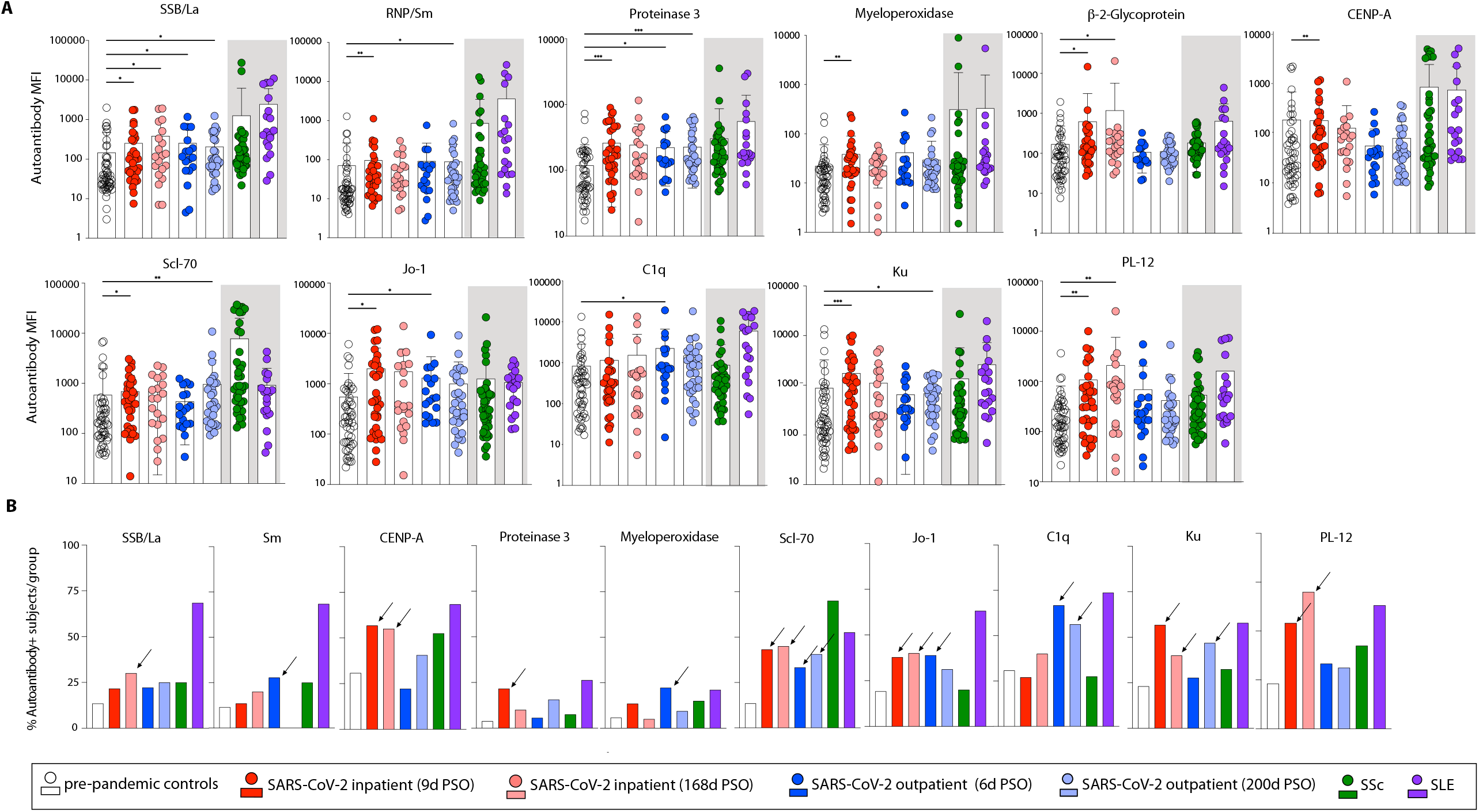
Autoantibodies associated with connective tissue autoimmune diseases and related conditions are found more frequently in SARS-CoV-2 infected individuals than pre-pandemic controls six months after symptom onset. (A) Autoantibody levels, shown as mean fluorescence intensity (MFI) were measured from six participant groups: (1) pre-pandemic controls (2) inpatient SARS-CoV-2+ with samples drawn ≤30 days (2) or ≥90 days (3) post symptom onset; outpatient SARS-CoV-2+ with samples drawn ≤30 days (4) or ≥90 days (5) post symptom onset; scleroderma (SSc) (5) and Systemic Lupus Erythromatasus (SLE) (6). Average number of days post symptom onset (PSO) for each SARS-CoV-2 sample group is shown. In (A), statistical comparisons (Kruskal-Wallis tests) were performed between pre-pandemic controls and the four SARS-CoV-2 infected sample groups only. *p<0.05, **p0.01, ***p0.001. Gray shading indicates exclusion from statistical comparisons. (B) autoantibody hits were defined using different MFI cutoffs for each readout (listed in Supplemental Figure 1) and the percentage of individuals with a hit for the autoantibody was determined for all groups. Arrows denote results 15% or higher than pre-pandemic controls. Of the 17 autoantibodies measured, only results with statistical significance are shown in (A) or with at least one of the SARS-CoV-2+ groups with >15% difference in frequency in (B) are shown; all other results are presented in Supplemental Figure 1.

We next determined the percentage of participants in each group with positive hits for each antibody (as described in Methods). Six (SSB/La, CENP-A, Scl-70, Jo-1, Ku, and PL-12) and three (SCL-70, C1q, and Ku) autoantibodies scored as positive hits in ≥15% of the SARS-CoV-2 inpatient and outpatient participants ≥90 days post symptom onset, respectively, compared with the controls (**Figure 1B**). Notably, the majority of SARS-CoV-2 inpatients possessed CENP-A and PL-12 autoantibodies at both acute and convalescent time points, and most SARS-CoV-2 outpatients possessed C1q autoantibodies at both time points at roughly twice the frequencies found in uninfected controls (**Figure 1B**). Scl-70 and Ku autoantibodies were present at ≥15% higher frequencies than pre-pandemic controls 90 days or greater post symptom onset among both the inpatient and outpatient participant groups (**Figure 1B**). Other autoantibodies measured were similar in the percentages of positive hits between the five sample groups (**Supplemental Figure 1B**). Taken together, these results demonstrate higher expression of autoantibodies of inpatient and outpatient SARS-CoV-2 infected individuals during both acute disease and six months after acute illness as compared with pre-pandemic controls. All MFI values for the samples included in these analyses are listed in **Supplemental Figure 2**.

### SARS-CoV-2 infection is an independent predictor of high autoantibody levels at acute and convalescent time points after adjusting for participant demographics and comorbidities

We next conducted univariate analysis comparing the differences between pre-pandemic and the SARS-CoV-2 infected groups, ≤30 days (average 11.6 days) and ≥90 days (average 188.4 days) post symptom onset (PSO) and multiple linear regressions were performed from the significantly different comparisons (p-values <0.05) between pre-pandemic and SARS-CoV-2 samples to adjust for the impact of demographics and comorbidities on the autoantibody differences noted (data not shown). At ≤30 days PSO, SARS-CoV-2 infection was revealed as an independent factor associated with elevated MFIs of four autoantibodies: (1) SSB/La (2.06-fold, p=0.009), (2) Proteinase 3 (2.70-fold, p<0.0001), (3) Jo-1 (4.06-fold, p<0.0001), and (4) Ku (2.36-fold, p=0.0018) (**Figure 2A**). At ≥90 days PSO, SARS-CoV-2 infection was associated with higher autoantibody MFIs for SSB/La (2.85-fold, p=0.0002), Sm (1.85-fold, p=0.0094) Proteinase 3 (2.78-fold, p<0.0001), Myeloperoxidase (1.6-fold, p=0.012), Jo-1 (2.48-fold, p=0.0023), and Ku (1.91-fold, p=0.0177) (**Figure 2B**). The other autoantibodies with higher average MFI in the SARS-CoV-2 infected groups compared to the pre-pandemic controls in univariate analysis were not significantly different after adjusting for demographics and comorbidities or, in some cases, the model assumptions did not satisfy with regression diagnostics (complete output tables can be found in **Supplemental Tables 1-4**).

**Figure 2.**
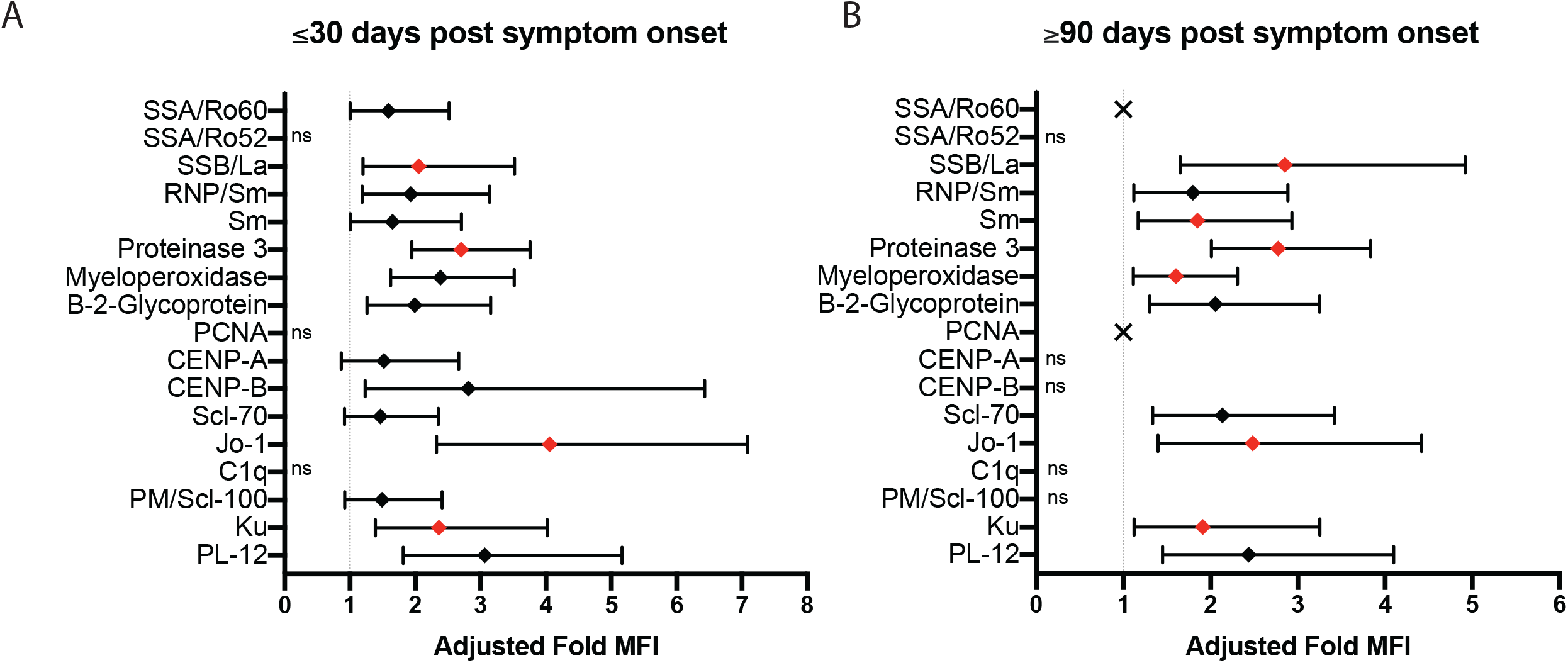
SARS-CoV-2 infection associated with elevated autoantibody levels at acute and convalescent timepoints compared to pre-pandemic controls in adjusted analysis. Multiple linear regression was performed to adjust for demographics and comorbidities between SARS-CoV-2 groups (A) ≤30 days post symptom onset (PSO) and (B) ≥90 days PSO compared to the pre-pandemic group. Data points represent MFI fold changes compared to the pre-pandemic. Red and black diamonds indicate statistically significant and not significant estimates (or unmet model requirements), respectively. “X” specifies SARS-CoV-2 infection did not associate with autoantibody level in adjusted analysis. “ns” indicates where univariate comparisons were not significant and multiple variable assessment was not conducted. Error bars represent 95% confidence intervals.

### SARS-CoV-2 infected individuals possess a higher number of autoantibodies than unexposed controls at both acute and convalescent time points

We next compared the average numbers of autoantibody hits per participant from the pre-pandemic and SARS-CoV-2 infected groups. In univariate analysis, pre-pandemic participants possessed a lower number of autoantibodies (2.9) compared to acutely infected SARS-CoV-2 participants (both inpatient and outpatient combined) (4.72 autoantibody hits, on average, 95% CI 3.826 to 5.607, p-value = 0.0059) and participants more than 90 days after symptom onset (4.54, autoantibody hits, on average, 95% CI 3.782 to 5.295, p-value = 0.0054) (**Figure 3**). Greater autoantibody hits per participant were found among hospitalized individuals both during acute illness and more than 90 days into convalescence (data not shown). Poisson multiple linear regression was performed to adjust for demographics and comorbidities, and this revealed that SARS-CoV-2 infection as an independent variable associated with approximately 1.6-fold greater autoantibody hits at both ≤30 days (<0.0001) and >90 days (p<0.0001) PSO (data not shown). In sum, SARS-CoV-2 infection is linked to a greater number of autoantibody hits per participant at both acute and convalescent stages compared to unexposed controls.

**Figure 3.**
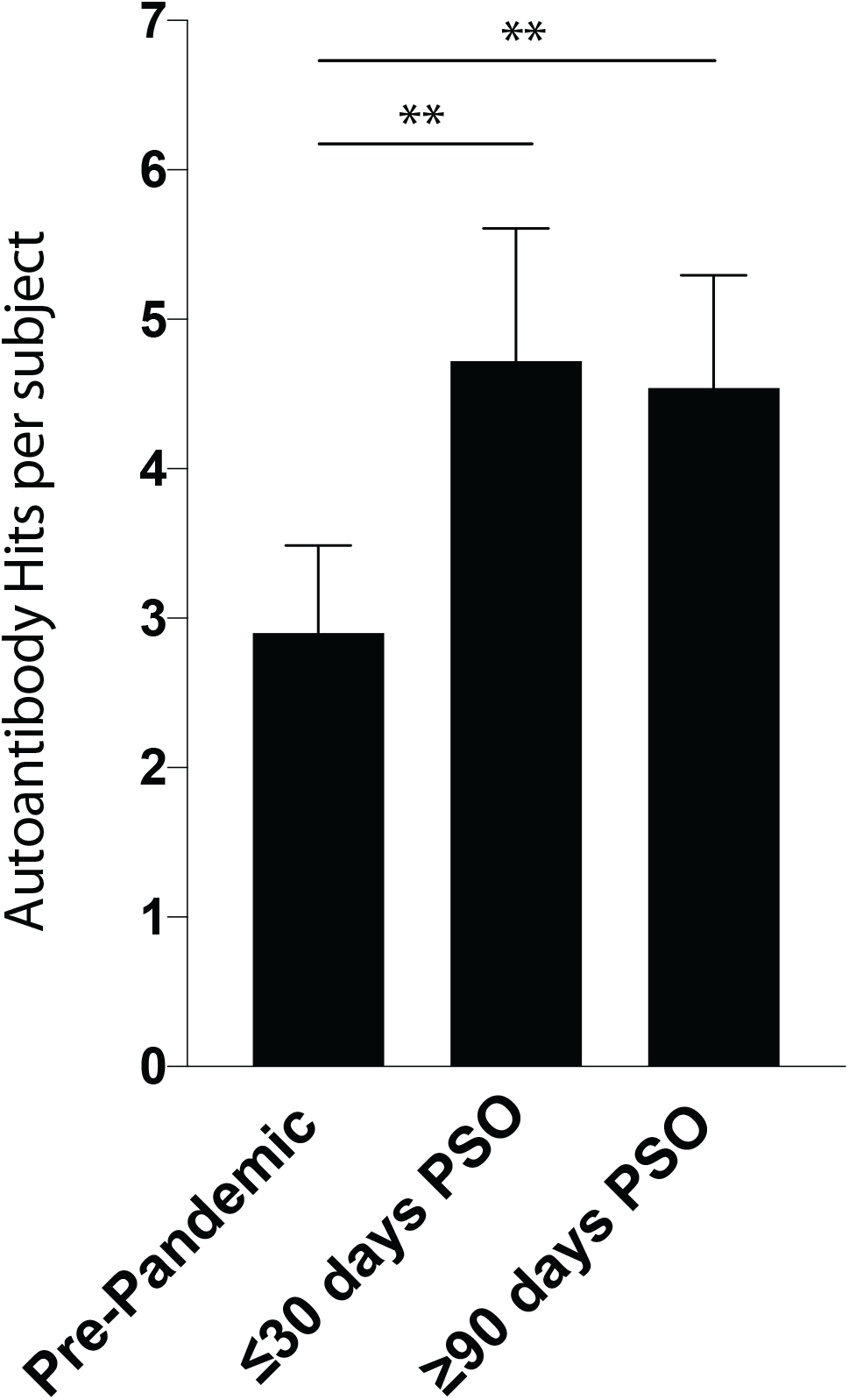
SARS-CoV-2 infected individuals possess a higher number of autoantibodies than uninfected controls during both acute illness and after months of convalescence. Univariate analysis showed SARS-CoV-2 participants (inpatient and outpatient) had higher number of autoantibodies positive at both acute and convalescent time points compared to pre-pandemic controls. ** indicates p value <0.01.

### Autoantibody signatures of individual participants six months after COVID-19 symptom onset resemble pre-pandemic controls and not individuals with SSc or SLE

We next investigated if the expression patterns of autoantibodies found in SARS-CoV-2 convalescent individuals (all ≥90 days PSO, average 185 days) resembled either of two clinically established autoimmune diseases, scleroderma (SSc) and systemic lupus erythematosus (SLE). We employed Partial Least Squares Determinant Analysis (PLSDA) to the 17 autoantibody MFIs measured from the following sample groups: (1) pre-pandemic, (2) SARS-CoV-2 infected convalescent (hospitalized and outpatient combined) with all samples collected ≥90 days after symptom onset, (3) SSc, and (4) SLE. PLSDA determines which measurements from each participant would “fit” that participant into its clinical cohort and verifies if such classification is statistically significant. As expected, the autoantibody signatures of the SSc, SLE, and pre-pandemic groups separated distinctly and with significance; however, the convalescent SARS-CoV-2 autoantibody signature was similar to the pre-pandemic participants and neither of the autoimmune diseased groups (**Figure 4**). Loadings on VD1 and VD2 show the specific autoantibodies that determined the differences between the groups, when found, with SSc and SLE groups distinctly defined via higher expression of many autoantibodies, the top five of which are of Sm, RNP/SM, C1q, Proteinase 3, and SSA/R060 (in this order) (**Supplemental Figure 3A**). Systemic sclerosis is defined from the other three groups most strongly by higher expression of CENP-A, CENP-B, and Scl-70 (**Supplemental Figure 3B**).

**Figure 4.**
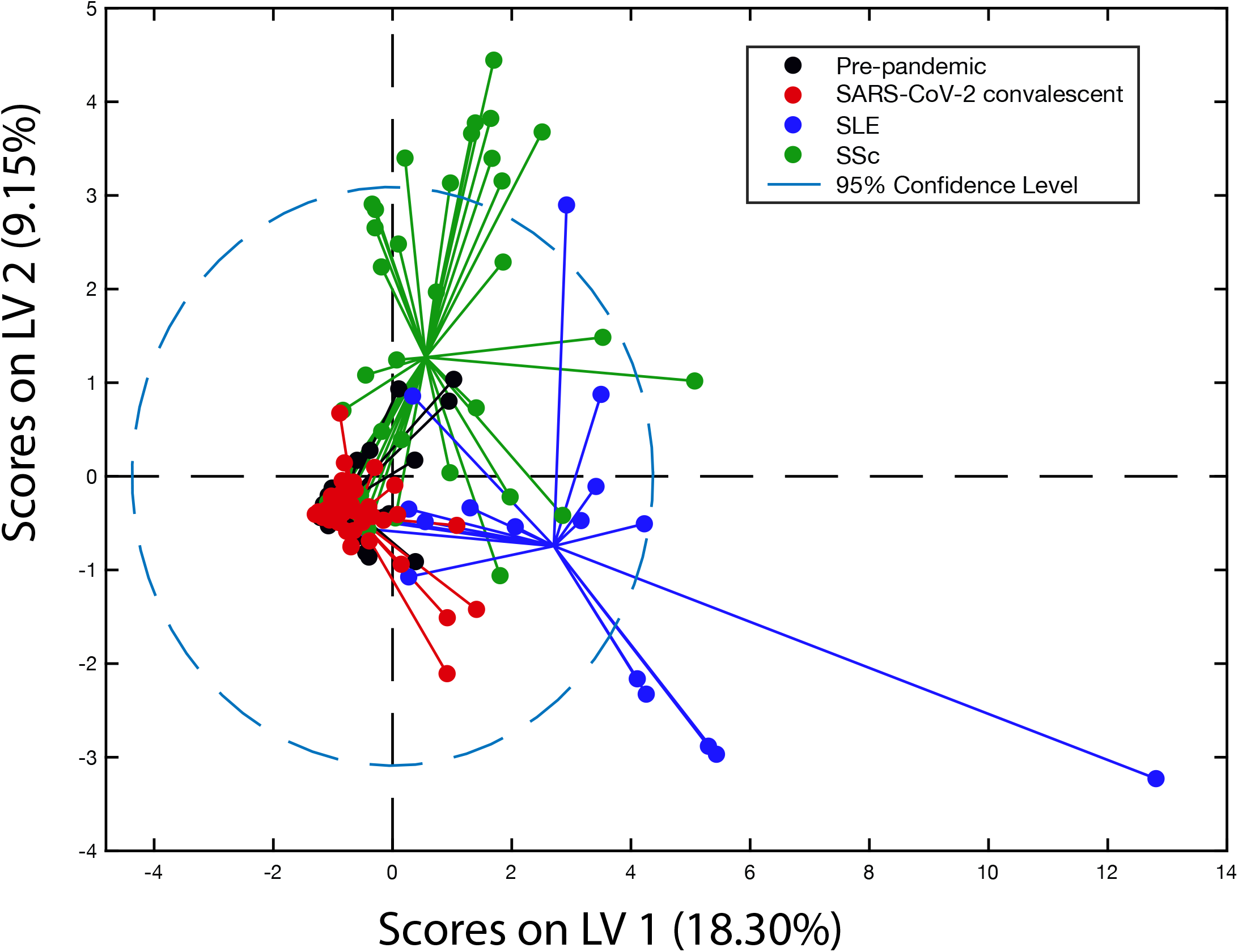
Partial Least Squares Discriminant Analysis (PLSDA) of autoimmune antibodies from pre-pandemic, convalescent SARS-CoV-2 infected, SLE and SSc individuals. A two-dimensional PLSDA model distinguished autoantibody profiles for pre-pandemic (black), convalescent SARS-CoV-2 infected (red), and SLE (blue) and SSc (green) individuals with >64% prediction confidence. Each data point represents scores generated by the model composed of 17 autoantibody measurments for a given individuals mapped into the two-dimersional latent variable space. The percentages on the axes show the percent variance in the datset captured by a particular LV. Dotted line shows the 95% confidence interval.

### A ‘negative, then positive’ expression pattern for one or more autoantibodies is present in 18% of outpatient and over 50% of hospitalized SARS-CoV-2 infected individuals

Autoantibody levels were followed over time from all SARS-CoV-2 infected individuals from symptom onset into months of convalescence (as described in Methods). Longitudinal changes in antibody levels were classified as one of four expression patterns: ‘positive for all time points’ (blue); ‘negative, then positive’ (red); ‘positive, then negative’ (purple); and ‘variable expression’ (green). These color-coded expression patterns are shown two ways: (1) MFI results of each autoantibody for all time points from the outpatient and inpatient individuals (**Supplemental Figures 4 and 5, respectively**); and (2) each participant’s composite results shown as color coded squares for all 17 autoantibodies measured (**Figure 5**). Of the 33 outpatients, six possessed at least one autoantibody with a ‘positive, then negative’ profile (red), and of these six individuals, three possessed two or more autoantibodies with this expression pattern, indicating putative induction of new immune responses to multiple self-antigens in these participants (**Figure 5B**). Autoantibodies with this expression pattern from multiple outpatients included anti-Ku (n=3), anti-Jo-1 (n=2) and anti-SSB-La (n=2). Strikingly, 53% of the hospitalized participants possess the ‘negative, then positive’ expression pattern for at least one autoantibody (**Figure 5B**). Also, of the inpatient participants with at least one autoantibody in this category, six exhibited three or more antibodies with this expression pattern (**Figure 5B**).

**Figure 5.**
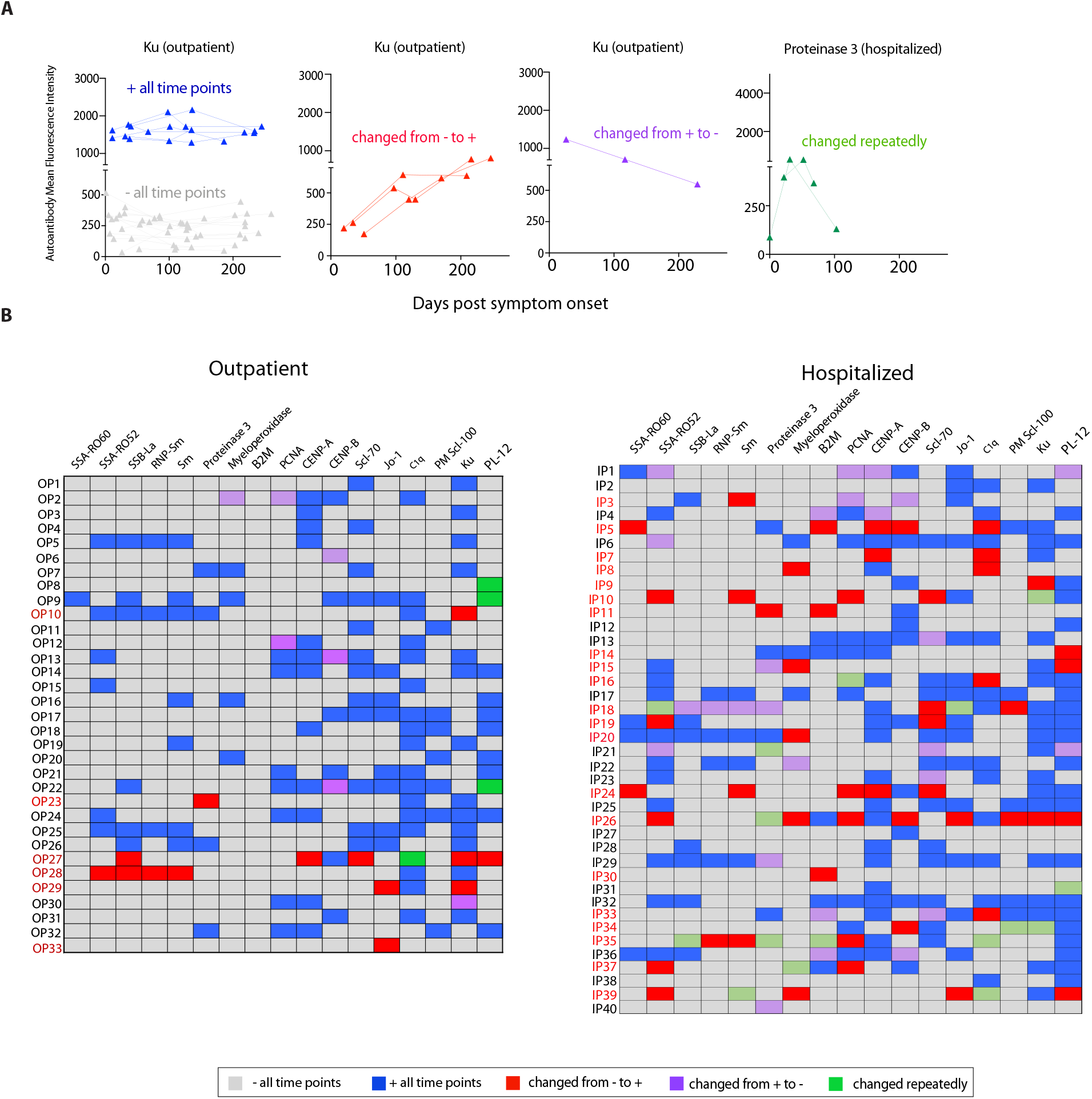
Longitudinal autoantibody expression in outpatient and hospitalized SARS-CoV-2 infected individuals reveals evidence of autoantibody induction with sustained expression. (A) Examples of longitudinal autoantibody expression patterns as defined. (B) Heat map showing the expression changes for all 17 autoantibodies measured for each participant. Individuals with an induction pattern of one or more autoantibodies is noted in red.

### A predominant autoantibody temporal expression pattern in both outpatient and hospitalized participants was positivity at all time points, indicating very early induction or pre-existence prior to viral exposure

Of all the autoantibody expression patterns scored that included at least one positive hit at one time point (blue, red, purple, and green colored results, **Figure 5**) many were found to be positive at all time points measured (**blue boxes, Figure 5B**). Given the relative stability of the MFI of these blue scored autoantibodies over time (**Supplemental Figures 4 and 5**), it is likely that these autoantibodies were present prior to SARS-CoV-2 infection.

## Discussion

Here, we demonstrate that autoantibodies linked to autoimmune connective tissue disorders are found at higher frequencies among SARS-CoV-2 infected individuals months into convalescence compared to pre-pandemic controls and exhibit temporal trajectories indicating initiation of new autoimmune responses. The induction of autoantibodies after viral infection is an established phenomenon [21–24], and our study is limited as it lacks comparative longitudinal data on the trajectory of autoantibodies from individuals infected with other pathogens. However, our results suggest that SARS-CoV-2 treads a sizeable autoimmune ‘footprint’ at least in the first six months after infection. We found notably higher autoantibodies in convalescent SARS-CoV-2 infected individuals in comparison with participant samples banked before the pandemic (**Figure 1**) and after adjusting for participant demographics and medical conditions, SARS-CoV-2 infection history significantly associated with the presence of many autoantibodies, even when this infection occurred an average of six months in the past (**Figure 2**). The pre-pandemic control group was comprised of US adult population with many decades of presumed microbial exposure. One can therefore surmise two possibilities to explain our findings: (1) overall, the cumulative impact of previous infections in the United States on middle aged adults yields lower autoantibody induction than SARS-CoV-2 infection; or (2), elevated autoantibodies six months post infection with many pathogens is common and this expression will wane in the longer term (years). To investigate the latter possibility, additional time points from many participants in this cohort will be analyzed to determine if the autoantibodies are still present 1.5 years after the initial infection.

Our longitudinal analysis shows evidence of induction and sustained expression of autoantibodies for six months in 18% of outpatient and 53% of hospitalized individuals infected with SARS-CoV-2 (**Figure 5**). Proposed mechanisms for the emergence of new autoantibodies during viral infections include molecular mimicry, epitope spreading, and bystander activation [25]. It is interesting to note that a wide panoply of self-epitopes are recognized by antibodies in severe COVID-19 [3, 26]; similarly, our results show that most of the autoantibodies we measured were higher in SARS-CoV-2 infected participants than controls (**Figure 1**). This wide breadth of antibody reactivities to self-epitopes may be a by-product of the immune cell homeostatic proliferation that occurs in response to the profound lymphopenia that is distinctly common with this infection [27–31]. Specifically, B and T cells with receptor specificities reactive to self-peptides that have escaped central tolerance may undergo misguided differentiation into effector cells from extensive cell turnover to fill empty immune space. Proliferation stimulated by homeostasis drives naïve T cells into memory cells without antigen present [32]; connections between lymphopenia and autoimmunity are well established [33–35] and COVID-19 severity positively correlates with both lymphopenia [29, 36] and evidence of novel autoantibodies in the circulation (**Figure 5**). Also, lymphopenia in the presence of massive inflammation results in T regulatory cell dysfunction [37], another likely driver of loss of immune tolerance. Future studies comparing plasma markers of inflammation, the magnitude and temporal kinetics of lymphopenia (including time to cell recovery), and autoantibody induction and expression patterns into SARS-CoV-2 convalescence would help determine if immune cell depletion then recovery drives autoimmunity in this infection.

Many SARS-CoV-2 infected participants in our study possessed autoantibodies with an expression pattern that suggests establishment very early post- or prior to SARS-CoV-2 exposure (‘positive all time points’) with levels that often remain stable months into convalescence (**Supplemental Figures 4, 5; Figure 5**). Anti-type I IFN antibodies, present early in SARS-CoV-2 infection, correlate with more severe COVID-19, suggesting a possible direct impact of these autoantibodies on the anti-viral immune response [1]. All individuals in our study were symptomatic, whether outpatient or hospitalized. It is possible that pre-existing autoantibodies are positively linked to a symptomatic outcome after SARS-CoV-2 exposure but a comparison of autoantibody expression in asymptomatic individuals is required. Two infections that are linked to self-reactive immune responses and/or can negatively impact immunity to new infections are Epstein Barr Virus (EBV) and Cytomegalovirus (CMV) [38–40]. Recently, a large study of following young adults in the military demonstrated a convincing link between history of EBV infection and eventual diagnosis of multiple sclerosis [41], with a further mechanistic study suggesting this may be caused due to molecular mimicry between EBV protein EBNA1 and GlialCAM expressed by oligodendrocytes [42]. Similar maladaptive responses to EBV infection have also been implicated in SSc and SLE cases [43, 44]; additionally, a recent cohort correlated EBV viremia with presence of some PASC symptoms [45]. The presence and/or level of control of EBV, CMV, and other chronic infections may directly impact the magnitude of an effective SARS-CoV-2 immune response and an investigation of possible connections between these chronic infections and COVID-19 severity is warranted.

Our multivariate PLSDA analysis found that the autoantibody signatures of SARS-CoV-2 infected participants resembled controls and not individuals with SSc or SLE, with the letter two groups distinct from one another and controls (**Figure 4**). The concept of infections leading to induction of a “fertile field” (increased propensity for acquiring new autoimmune diseases upon an additional environmental triggers) [46] may also be applicable here. Also, these PLSDA results indicate that SARS-CoV-2 convalescent participants undergo a disorganized, random autoantibody induction, lending further support to the postulated mechanisms described above (lymphopenia-induced loss of tolerance) and distinct from what would occur with molecular mimicry or epitope spreading. Importantly, the presence of individual autoantibodies has been shown to be a preclinical signal for eventual development of an eventual autoimmune disorder [47] and it is well established that positivity of individual autoantibodies precedes presentation of clinical disease, often by many years [48–52]. Also, the presence of SLE and SSc related autoantibodies was a strong predictor of certain PASC phenotypes two-three months after recovery from acute illness [45]. Examining autoantibody signatures for organization into patterns such as those found in SSc and SLE participants in conjunction with clinical outcomes such as incidence of autoinflammatory and autoimmune disorders can help elucidate the pathophysiology of these conditions.

There are additional limitations to our study. We sought to understand the immunologic trajectories noted in patients with this novel pathogen over longer periods post exposure. However, our study is not linked to clinical outcomes of interest during convalescence but rather is providing a broad background understanding of autoantibody dynamics in SARS-CoV-2 convalescent individuals. We also only examine a limited number of autoantibodies generally linked to autoimmune connective tissue disorders, the symptoms of which have been suggested to be consistent with PASC. To date, one of the issues to linking clinical outcomes such as PASC to immunological phenomenon has been the lack of a unifying clinical case criteria and understanding of the diverse phenotypes for this condition. The National Institutes of Health recently funded a large prospective cohort with the goal of understanding post-acute sequalae of SARS-CoV-2, entitled Researching COVID to Enhance Recovery (RECOVER), which may make such investigations possible in near future (https://recovercovid.org). Additionally, as mentioned above, our study does not have similar longitudinal samples from other viral infections, or for pre-pandemic controls (particularly those that might be hospitalized with severe illness). These types of controls should be included in future prospective cohorts of SARS-CoV-2 infection. We also do not have pre-infection samples for the SARS-CoV-2 cases, which would enable discerning early induced from pre-existing autoantibodies.

The timing of sample collection for this study was early in the pandemic (spring and summer of 2020), when there were not vaccines available which would impact the initial immunological response; also, there were very low levels of seasonal coronaviruses and other common pathogens in circulation due to lockdowns and other public health restrictions. This is important to consider as therefore our findings are likely reflective of a more naïve response to SARS-CoV-2 and the autoantibody changes noted over time are less likely to be due to other infections than under normal circumstances. Future longitudinal studies of individuals infected with SARS-CoV-2 far later in the pandemic, with a mix of vaccination and SARS-CoV-2 infection histories, will be informative to gain new insight into how different signatures of virus-specific pre-existing immunity and concurrent infections impacts loss of immunological self-tolerance.

At this writing, over 513 million cases of SARS-CoV-2 infection have been recorded globally with the real estimates expected to be twice or thrice that number due to under-reporting and under-diagnosis. In the wake of this viral pandemic are reports of a panoply of symptoms that are not resolving in all individuals after acute illness and new symptomologies arising weeks-months after initial infection, including among patients who are initially asymptomatic. Elucidating the interplay between factors such as pre-existing infections and immune memory, acute viral damage and inflammation, lymphopenia, and new onset immune responses is imperative to combat the mortality and morbidity from SARS-CoV-2 and likely many other infections as well.

## Supporting information

Supplemental Data and Graphs

## Data Availability

All data produced in the present work are contained in the manuscript

## Figure Legends

**Supplemental Figure 1**. Autoantibody Mean Fluorescence Intensity (MFI) (A) and percentage of autoantibody positive individuals within the six participant groups (B); for all data shown there was statistical significance between the pre-pandemic and SARS-CoV-2 infected groups.

**Supplemental Figure 2:** MFI values for autoantibodies measured from all individuals. Positive ‘hits’ for each result are noted in pink.

**Supplemental Figure 3**. Bar plots showing the loadings on LV1 (A) and LV2 (B) of 17 autoantibodies measurments used to train the PLSDA model. LV1 is the latent variable that predominantely separated the SLE individuals from the other clinical groups. LV2 is he latent variable that predominantely separated the SSc individuals from the other clinical groups. The Y-axis quantifies the positive or negative contribution of a particular parameter to the inducated LV. The colored dots denote the autoantibodies with a VIP score greater than 1 (statistically significant) for each clinical group in the model. Hatched bars indicate VIP>1 scores for the SLE group in panel A and VIP >1 scores for the SSc group in panel B.

**Supplemental Figure 4**. Raw MFI data of longitudinal autoantibody results from all outpatient SARS-CoV-2 individuals. Different defined patterns of expression (as described in Figure 5) are noted by the different colors.

**Supplemental Figure 5**. Raw data of longitudinal autoantibody results from all hospitalized SARS-CoV-2 individuals. Different defined patterns of expression (as described in Figure 5) are noted by the different colors.

