## Supplemental Data and Graphs for "Longitudinal analysis reveals elevation then sustained higher expression of autoantibodies for six months after SARS-CoV-2 infection"

**Supplemental Table 1.** Multiple Linear Regression: ≤30 days post-SARS-CoV-2 symptom onset

| AAB | Variable | Estimate | 95% CI (asymptotic) | P value | P value summary |
|---|---|---|---|---|---|
| **SSA/Ro60** | | | | | |
| β0 | Intercept | 1.68 | 1.486 to 1.875 | <0.0001 | **** |
| β1 | SARS-CoV-2 | 0.2011 | 0.0009555 to 0.4013 | 0.0489 | * |
| β2 | Sex: Female | -0.2313 | -0.4473 to -0.01525 | 0.0361 | * |
| β3 | Race: White | -0.1901 | -0.3949 to 0.01466 | 0.0684 | ns |
| β4 | Ethnicity: Hispanic/Latino | -0.2445 | -0.5428 to 0.05381 | 0.1071 | ns |
| β5 | Lung Disease | 0.2659 | 0.02635 to 0.5055 | 0.03 | * |
| β6 | Immunosuppressed | -0.2912 | -0.6177 to 0.03527 | 0.0798 | ns |
| **SSB/La** | | | | | |
| β0 | Intercept | 1.668 | 1.505 to 1.831 | <0.0001 | **** |
| β1 | SARS-CoV-2 | 0.3131 | 0.07997 to 0.5463 | 0.009 | ** |
| β2 | Heart Disease | 0.3233 | 0.01018 to 0.6365 | 0.0431 | * |
| β3 | Immunosuppressed | -0.2845 | -0.6659 to 0.09689 | 0.142 | ns |
| **RNP/Sm** | | | | | |
| β0 | Intercept | 1.305 | 1.157 to 1.452 | <0.0001 | **** |
| β1 | SARS-CoV-2 | 0.2857 | 0.07484 to 0.4966 | 0.0084 | ** |
| β2 | Heart Disease | 0.265 | -0.01820 to 0.5482 | 0.0663 | ns |
| β3 | Immunosuppressed | -0.4536 | -0.7984 to -0.1087 | 0.0105 | * |
| **Sm** | | | | | |
| β0 | Intercept | 2.056 | 1.902 to 2.209 | <0.0001 | **** |
| β1 | SARS-CoV-2 | 0.2178 | 0.003095 to 0.4325 | 0.0468 | * |
| β2 | Heart Disease | 0.3153 | 0.02574 to 0.6049 | 0.0331 | * |
| β3 | Lung Disease | 0.1824 | -0.06693 to 0.4317 | 0.1498 | ns |
| β4 | Immunosuppressed | -0.5201 | -0.8721 to -0.1681 | 0.0042 | ** |
| **Proteinase 3** | | | | | |
| β0 | Intercept | 2.019 | 1.774 to 2.263 | <0.0001 | **** |
| β1 | SARS-CoV-2 | 0.432 | 0.2892 to 0.5748 | <0.0001 | **** |
| β2 | Age | -0.00364 | -0.007816 to 0.0005340 | 0.0867 | ns |
| β3 | Sex: Female | -0.1587 | -0.2993 to -0.01798 | 0.0275 | * |
| β4 | Race: Black | 0.1851 | 0.04975 to 0.3204 | 0.0079 | ** |
| β5 | HIV | 0.2106 | 0.02374 to 0.3975 | 0.0276 | * |
| β6 | Immunosuppressed | -0.2112 | -0.4285 to 0.006177 | 0.0567 | ns |
| **Myeloperoxidase** | | | | | |
| β0 | Intercept | 0.9201 | 0.5877 to 1.252 | <0.0001 | **** |
| β1 | SARS-CoV-2 | 0.378 | 0.2100 to 0.5461 | <0.0001 | **** |
| β2 | Age | 0.004616 | -0.0008467 to 0.01008 | 0.0967 | ns |
| β3 | Sex: Female | -0.167 | -0.3482 to 0.01424 | 0.0705 | ns |
| β4 | Race: White | -0.1576 | -0.3229 to 0.007706 | 0.0614 | ns |
| β5 | Lung Disease | 0.296 | 0.09918 to 0.4928 | 0.0036 | ** |
| β6 | Autoimmune | 0.3435 | 0.1323 to 0.5547 | 0.0017 | ** |
| β7 | Immunosuppressed | -0.4635 | -0.7245 to -0.2024 | 0.0007 | *** |
| β8 | Hypertension | -0.1903 | -0.3896 to 0.009076 | 0.0612 | ns |
| **B-2-Glycoprotein** | | | | | |
| β0 | Intercept | 1.492 | 1.150 to 1.835 | <0.0001 | **** |
| β1 | SARS-CoV-2 | 0.2997 | 0.1007 to 0.4987 | 0.0035 | ** |
| β2 | Age | 0.00586 | 2.735e-005 to 0.01169 | 0.049 | * |
| β3 | HIV | 0.2454 | -0.01860 to 0.5094 | 0.0681 | ns |
| **CENP-A** | | | | | |
| β0 | Intercept | 2.612 | 2.389 to 2.836 | <0.0001 | **** |
| β1 | SARS-CoV-2 | 0.1824 | -0.06095 to 0.4257 | 0.1401 | ns |
| β2 | Race: White | -0.2612 | -0.5029 to -0.01959 | 0.0344 | * |
| β3 | Heart Disease | 0.3788 | 0.05359 to 0.7040 | 0.0229 | * |
| β4 | Autoimmune | 0.2852 | -0.01848 to 0.5889 | 0.0654 | ns |
| β5 | Immunosuppressed | -0.367 | -0.7588 to 0.02476 | 0.066 | ns |
| **CENP-B** | | | | | |
| β0 | Intercept | 1.526 | 1.234 to 1.818 | <0.0001 | **** |
| β1 | SARS-CoV-2 | 0.4495 | 0.09084 to 0.8082 | 0.0146 | * |
| β2 | Sex: Female | -0.3555 | -0.7202 to 0.009142 | 0.0559 | ns |
| β3 | Ethnicity: Hispanic/Latino | 0.3737 | -0.1114 to 0.8587 | 0.1295 | ns |
| β4 | HIV | 0.6565 | 0.1900 to 1.123 | 0.0063 | ** |
| β5 | Lung Disease | 0.4236 | 0.02204 to 0.8251 | 0.0389 | * |
| β6 | Immunosuppressed | -0.376 | -0.9135 to 0.1614 | 0.1682 | ns |
| **Scl--70** | | | | | |
| β0 | Intercept | 2.202 | 2.047 to 2.357 | <0.0001 | **** |
| β1 | SARS-CoV-2 | 0.1669 | -0.03804 to 0.3717 | 0.1093 | ns |
| β2 | Ethnicity: Hispanic/Latino | 0.229 | -0.06529 to 0.5233 | 0.1258 | ns |
| β3 | Hypertension | 0.2405 | 0.03018 to 0.4508 | 0.0254 | * |
| **Jo-1** | | | | | |
| β0 | Intercept | 2.099 | 1.902 to 2.296 | <0.0001 | **** |
| β1 | SARS-CoV-2 | 0.6084 | 0.3663 to 0.8505 | <0.0001 | **** |
| β2 | HIV | 0.5682 | 0.2554 to 0.8811 | 0.0005 | *** |
| β3 | DM | -0.3907 | -0.7023 to -0.07907 | 0.0145 | * |
| β4 | Heart Disease | 0.253 | -0.07959 to 0.5857 | 0.1343 | ns |

| | | | | | |
|---|---|---|---|---|---|
| β5 | Lung Disease | 0.2655 | 0.0009944 to 0.5300 | 0.0492 | * |
| β6 | Immunosuppressed | -0.7157 | -1.088 to -0.3430 | 0.0002 | *** |
| β7 | Hypertension | 0.2666 | -0.01109 to 0.5442 | 0.0597 | ns |
| **Ku** | | | | | |
| β0 | Intercept | 2.286 | 2.115 to 2.456 | <0.0001 | **** |
| β1 | SARS-CoV-2 | 0.3736 | 0.1430 to 0.6042 | 0.0018 | ** |
| β2 | Immunosuppressed | -0.5398 | -0.9009 to -0.1788 | 0.0038 | ** |
| β3 | Hypertension | 0.3577 | 0.1186 to 0.5968 | 0.0037 | ** |
| **PL-12** | | | | | |
| β0 | Intercept | 2.088 | 1.906 to 2.271 | <0.0001 | **** |
| β1 | SARS-CoV-2 | 0.4864 | 0.2596 to 0.7132 | <0.0001 | **** |
| β2 | Sex: Female | -0.3525 | -0.5823 to -0.1226 | 0.003 | ** |
| β3 | HIV | 0.4416 | 0.1440 to 0.7392 | 0.004 | ** |
| β4 | Lung Disease | 0.2938 | 0.03958 to 0.5479 | 0.0239 | * |

**Supplemental Table 2.** Multiple Linear Regression: ≥90 days post-SARS-CoV-2 symptom onset

| AAB | Variable | Estimate | 95% CI (asymptotic) | P value | P value summary |
|---|---|---|---|---|---|
| **SSB/La** | | | | | |
| β0 | Intercept | 1.286 | 0.8832 to 1.688 | <0.0001 | **** |
| β1 | SARS-CoV-2 | 0.4551 | 0.2182 to 0.6920 | 0.0002 | *** |
| β2 | Race: White | 0.379 | -0.003725 to 0.7617 | 0.0522 | ns |
| β3 | Race: Black | 0.4995 | 0.07138 to 0.9276 | 0.0227 | * |
| β4 | DM | 0.3125 | -0.03056 to 0.6556 | 0.0737 | ns |
| β5 | Hypertension | -0.2559 | -0.5504 to 0.03854 | 0.0877 | ns |
| **RNP/Sm** | | | | | |
| β0 | Intercept | 1.544 | 1.185 to 1.902 | <0.0001 | **** |
| β1 | SARS-CoV-2 | 0.2544 | 0.04861 to 0.4602 | 0.0159 | * |
| β2 | Age | -0.00458 | -0.01085 to 0.001693 | 0.1507 | ns |
| **Sm** | | | | | |
| β0 | Intercept | 2.088 | 1.946 to 2.229 | <0.0001 | **** |
| β1 | SARS-CoV-2 | 0.2672 | 0.06707 to 0.4673 | 0.0094 | ** |
| **Proteinase 3** | | | | | |
| β0 | Intercept | 1.988 | 1.855 to 2.122 | <0.0001 | **** |
| β1 | SARS-CoV-2 | 0.4433 | 0.3031 to 0.5836 | <0.0001 | **** |
| β2 | Sex: Female | -0.1384 | -0.2741 to -0.002598 | 0.0459 | * |
| β3 | Race: White | -0.1568 | -0.2912 to -0.02233 | 0.0227 | * |
| β4 | Autoimmune | 0.2258 | 0.05608 to 0.3955 | 0.0096 | ** |
| β5 | Immunosuppressed | -0.2659 | -0.4806 to -0.05128 | 0.0157 | * |
| **Myeloperoxidase** | | | | | |
| β0 | Intercept | 1.064 | 0.9313 to 1.196 | <0.0001 | **** |
| β1 | SARS-CoV-2 | 0.2047 | 0.04608 to 0.3633 | 0.012 | * |
| β2 | Race: Black | 0.2063 | 0.02179 to 0.3909 | 0.0288 | * |
| β3 | Hypertension | -0.189 | -0.3749 to -0.003120 | 0.0463 | * |
| **B-2-Glycoprotein** | | | | | |
| β0 | Intercept | 1.489 | 1.150 to 1.828 | <0.0001 | **** |
| β1 | SARS-CoV-2 | 0.313 | 0.1142 to 0.5119 | 0.0023 | ** |
| β2 | Age | 0.007024 | 0.0008989 to 0.01315 | 0.025 | * |
| β3 | HIV | 0.2434 | -0.01196 to 0.4988 | 0.0615 | ns |
| β4 | Heart Disease | -0.2666 | -0.5333 to 0.0001302 | 0.0501 | ns |
| β5 | Lung Disease | -0.1793 | -0.4331 to 0.07456 | 0.1642 | ns |
| **PCNA** | | | | | |
| β0 | Intercept | 2.527 | 2.281 to 2.772 | <0.0001 | **** |
| β1 | Race: White | -0.3416 | -0.6084 to -0.07482 | 0.0126 | * |
| β2 | HIV | -0.271 | -0.6189 to 0.07695 | 0.1254 | ns |
| β3 | Lung Disease | -0.2565 | -0.5996 to 0.08668 | 0.1412 | ns |
| β4 | Autoimmune | -0.378 | -0.7094 to -0.04654 | 0.0258 | * |
| **Scl-70** | | | | | |
| β0 | Intercept | 2.28 | 2.119 to 2.442 | <0.0001 | **** |
| β1 | SARS-CoV-2 | 0.3295 | 0.1251 to 0.5339 | 0.0019 | ** |
| β2 | Lung Disease | -0.2229 | -0.4982 to 0.05242 | 0.1114 | ns |
| β3 | Hypertension | 0.1747 | -0.06295 to 0.4123 | 0.1479 | ns |
| **Jo-1** | | | | | |
| β0 | Intercept | 2.145 | 1.933 to 2.356 | <0.0001 | **** |
| β1 | SARS-CoV-2 | 0.395 | 0.1446 to 0.6453 | 0.0023 | ** |
| β2 | Ethnicity: Hispanic/Latino | 0.2746 | -0.06032 to 0.6096 | 0.1069 | ns |
| β3 | HIV | 0.2711 | -0.05906 to 0.6012 | 0.1064 | ns |
| β4 | Hypertension | 0.2224 | -0.04317 to 0.4880 | 0.0997 | ns |

| | | | | | |
|---|---|---|---|---|---|
| **Ku** | | | | | |
| β0 | Intercept | 2.494 | 2.241 to 2.746 | <0.0001 | **** |
| β1 | SARS-CoV-2 | 0.2811 | 0.04983 to 0.5124 | 0.0177 | * |
| β2 | Race: White | -0.1801 | -0.4131 to 0.05294 | 0.1283 | ns |
| β3 | HIV | -0.2162 | -0.5259 to 0.09346 | 0.169 | ns |
| β4 | Lung Disease | -0.2733 | -0.5639 to 0.01735 | 0.065 | ns |
| β5 | Hypertension | 0.2398 | -0.02294 to 0.5025 | 0.0732 | ns |
| **PL-12** | | | | | |
| β0 | Intercept | 2.446 | 2.082 to 2.809 | <0.0001 | **** |
| β1 | SARS-CoV-2 | 0.3867 | 0.1606 to 0.6128 | 0.001 | *** |
| β2 | Age | -0.00584 | -0.01196 to 0.0002785 | 0.0612 | ns |
| β3 | Sex: Female | -0.1595 | -0.3722 to 0.05321 | 0.14 | ns |
| β4 | HIV | 0.2576 | -0.02764 to 0.5427 | 0.0762 | ns |

**Supplemental Table 3.** Poisson multiple linear regression of ≤30 days PSO and pre-pandemic samples

| Parameter estimates | Variable | Estimate | 95% CI (profile likelihood) | P value |
|---|---|---|---|---|
| β0 | Intercept | 2.977 | 2.503 to 3.541 | <0.0001 |
| β1 | SARS-CoV-2 | 1.551 | 1.254 to 1.919 | <0.0001 |
| β2 | Female sex | 0.813 | 0.647 to 1.022 | 0.0761 |
| β3 | Heart disease | 1.675 | 1.315 to 2.134 | <0.0001 |
| β4 | Immunosuppression | 0.604 | 0.418 to 0.872 | 0.0072 |

**Supplemental Table 4.** Poisson multiple Linear Regression of ≥90 days PSO and pre-pandemic individuals

| Parameter estimates | Variable | Estimate | 95% CI (profile likelihood) | P value |
|---|---|---|---|---|
| β0 | Intercept | 2.798 | 2.300 to 3.404 | <0.0001 |
| β1 | SARS-CoV-2 | 1.662 | 1.342 to 2.059 | <0.0001 |
| β2 | Black race | 1.250 | 0.998 to 1.566 | 0.0526 |
| β3 | Lung disease | 0.631 | 0.464 to 0.860 | 0.0035 |

Supplemental Figure 1.

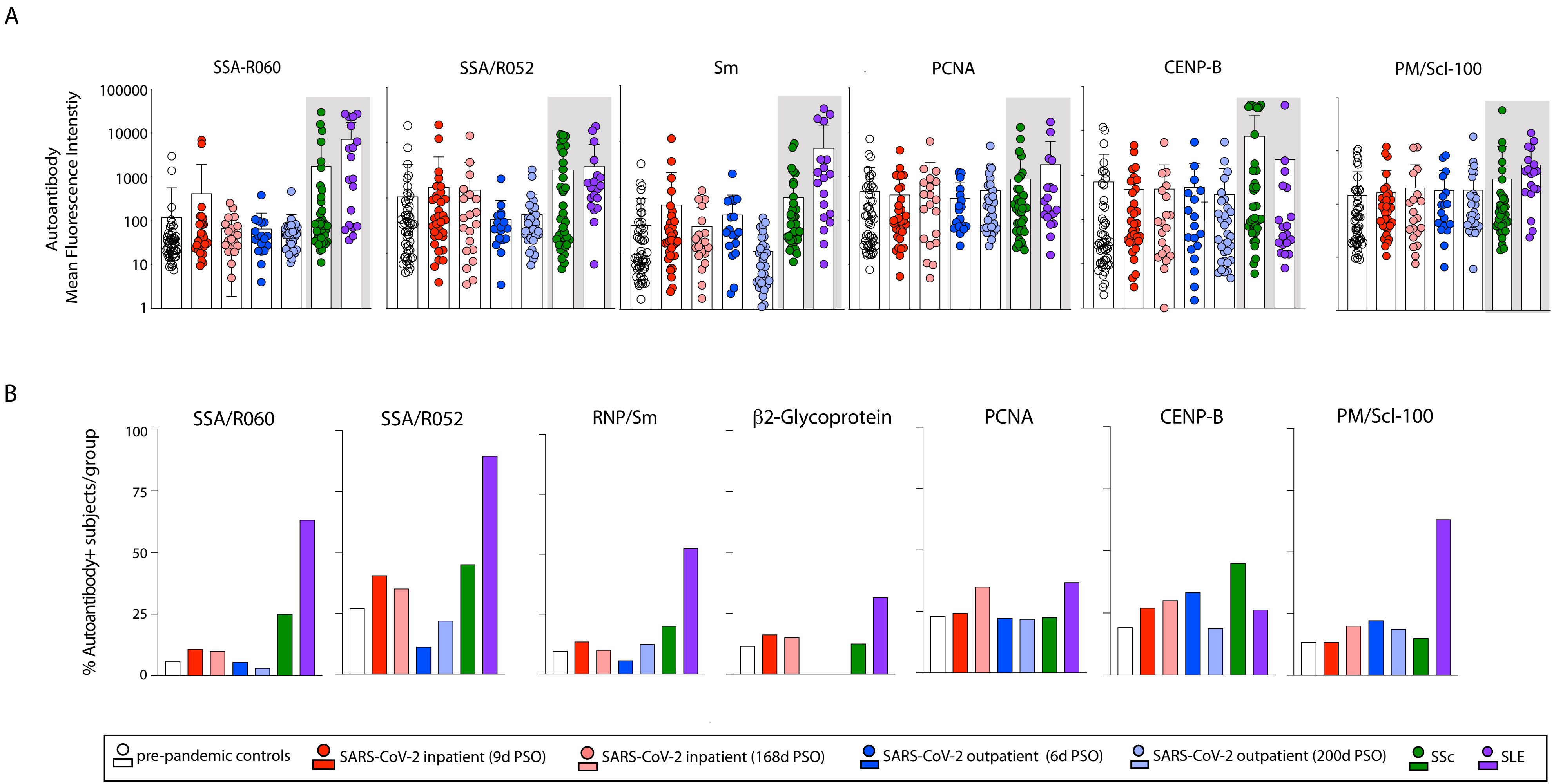

### Supplemental Figure 2

| Autoantibody | SSA/Ro60 | SSA/R052 | SSB/La | RNP/SM | Sm | Proteinase 3 | Myeloperoxidase | B2-Glycoprotein | PCNA | CENP-A | CENP-B | Scl-70 | Jo-1 | C1q | PM/Scl-100 | Ku | PL-12 |
|---|---|---|---|---|---|---|---|---|---|---|---|---|---|---|---|---|---|
| **MFI Cutoff** | **175** | **551** | **300** | **185** | **610** | **450** | **50** | **325** | **500** | **525** | **275** | **500** | **750** | **500** | **200** | **580** | **310** |
| ***Prepandemics*** | | | | | | | | | | | | | | | | | |
| **PP01** | 1394 | 1116 | 15 | 5 | 27 | 45 | 8 | 22 | 1031 | 113 | 7 | 233 | 271 | 57 | 184 | 209 | 182 |
| **PP02** | 15 | 429 | 26 | 10 | 72 | 207 | 10 | 237 | 1101 | 213 | 10298 | 407 | 444 | 70 | 1083 | 493 | 331 |
| **PP03** | 41 | 4110 | 81 | 45 | 241 | 58 | 28 | 52 | 213 | 20183 | 1911 | 6858 | 278 | 882 | 1010 | 10300 | 3632 |
| **PP04** | 24 | 245 | 18 | 6 | 109 | 193 | 166 | 40 | 17 | 313 | 1280 | 73 | 33 | 17 | 11 | 159 | 49 |
| **PP05** | 81 | 111 | 23 | 9 | 78 | 61 | 21 | 66 | 104 | 202 | 92 | 147 | 381 | 28 | 185 | 1081 | 176 |
| **PP06** | 36 | 8103 | 50 | 27 | 182 | 201 | 21 | 23 | 35 | 19519 | 836 | 522 | 297 | 347 | 68 | 875 | 146 |
| **PP07** | 34 | 1101 | 39 | 11 | 34 | 198 | 25 | 82 | 32 | 1574 | 46 | 147 | 172 | 47 | 35 | 145 | 175 |
| **PP08** | 36 | 217 | 76 | 26 | 356 | 57 | 19 | 85 | 23 | 284 | 104 | 104 | 59 | 24 | 58 | 121 | 122 |
| **PP09** | 36 | 709 | 18 | 5 | 54 | 35 | 5 | 356 | 17 | 45 | 11 | 163 | 35 | 1549 | 10 | 84 | 196 |
| **PP10** | 17 | 115 | 6 | 51 | 50 | 24 | 8 | 52 | 19 | 143 | 2 | 301 | 64 | 45 | 195 | 130 | 76 |
| **PP11** | 21 | 80 | 20 | 8 | 84 | 24 | 36 | 26 | 555 | 556 | 189 | 109 | 785 | 96 | 18 | 2630 | 59 |
| **PP12** | 21 | 163 | 60 | 32 | 263 | 29 | 13 | 97 | 3160 | 126 | 11 | 55 | 3195 | 173 | 22 | 36 | 62 |
| **PP13** | 47 | 493 | 24 | 17 | 269 | 35 | 8 | 8 | 388 | 44 | 10 | 426 | 69 | 5570 | 744 | 28 | 45 |
| **PP14** | 40 | 85 | 28 | 17 | 81 | 159 | 10 | 45 | 35 | 160 | 27 | 322 | 1018 | 342 | 137 | 220 | 247 |
| **PP15** | 31 | 88 | 584 | 10 | 59 | 35 | 3 | 12 | 934 | 53 | 4 | 94 | 29 | 107 | 58 | 164 | 66 |
| **PP16** | 16 | 65 | 17 | 9 | 61 | 56 | 10 | 122 | 46 | 183 | 1102 | 2130 | 162 | 569 | 38 | 586 | 435 |
| **PP17** | 41 | 891 | 24 | 19 | 196 | 176 | 14 | 110 | 278 | 203 | 597 | 181 | 347 | 1375 | 20 | 179 | 360 |
| **PP18** | 204 | 349 | 223 | 80 | 531 | 85 | 23 | 87 | 130 | 1704 | 648 | 6536 | 366 | 13272 | 49 | 229 | 304 |
| **PP19** | 15 | 454 | 30 | 12 | 40 | 98 | 20 | 311 | 81 | 270 | 1230 | 88 | 2332 | 233 | 35 | 89 | 554 |
| **PP20** | 21 | 201 | 30 | 16 | 70 | 73 | 22 | 248 | 40 | 385 | 32 | 106 | 143 | 114 | 556 | 716 | 89 |
| **PP21** | 53 | 62 | 17 | 10 | 174 | 143 | 15 | 67 | 619 | 619 | 5138 | 82 | 48 | 627 | 19 | 91 | 152 |
| **PP22** | 8 | 45 | 3 | 4 | 15 | 17 | 3 | 283 | 17 | 787 | 29 | 42 | 686 | 459 | 10 | 119 | 39 |
| **PP23** | 11 | 54 | 108 | 8 | 29 | 28 | 9 | 1003 | 2763 | 686 | 272 | 84 | 29 | 57 | 27 | 52 | 55 |
| **PP24** | 47 | 349 | 17 | 14 | 165 | 61 | 4 | 27 | 502 | 38 | 3 | 94 | 22 | 163 | 57 | 68 | 168 |
| **PP25** | 15 | 1622 | 215 | 11 | 29 | 62 | 17 | 69 | 28 | 3725 | 22 | 52 | 71 | 49 | 104 | 44 | 126 |
| **PP26** | 37 | 786 | 39 | 25 | 103 | 43 | 15 | 152 | 26 | 18091 | 26 | 80 | 69 | 327 | 829 | 13335 | 390 |
| **PP27** | 35 | 20409 | 689 | 465 | 1608 | 104 | 26 | 127 | 1396 | 702 | 31 | 220 | 437 | 693 | 318 | 1913 | 663 |
| **PP28** | 43 | 73 | 26 | 18 | 72 | 53 | 18 | 28 | 27 | 94 | 10 | 1452 | 29 | 163 | 116 | 73 | 470 |
| **PP29** | 18 | 149 | 40 | 17 | 86 | 183 | 74 | 65 | 30 | 1064 | 12100 | 127 | 529 | 2623 | 31 | 414 | 124 |
| **PP30** | 106 | 382 | 116 | 68 | 538 | 86 | 19 | 83 | 109 | 451 | 27 | 250 | 217 | 3974 | 603 | 751 | 1185 |
| **PP31** | 16 | 64 | 47 | 18 | 225 | 32 | 7 | 16 | 8 | 167 | 8 | 94 | 37 | 21 | 21 | 21 | 37 |
| **PP32** | 23 | 2292 | 1993 | 1289 | 3976 | 166 | 31 | 86 | 381 | 2298 | 206 | 398 | 796 | 1263 | 79 | 509 | 406 |
| **PP33** | 9 | 408 | 71 | 10 | 76 | 197 | 11 | 21 | 17 | 139 | 10 | 48 | 53 | 590 | 19 | 357 | 22 |
| **PP34** | 30 | 291 | 21 | 18 | 95 | 577 | 9 | 36 | 555 | 2279 | 36 | 182 | 140 | 638 | 22 | 39 | 67 |
| **PP35** | 21 | 495 | 303 | 187 | 861 | 48 | 20 | 27 | 44 | 2932 | 25 | 258 | 170 | 422 | 86 | 291 | 179 |
| **PP36** | 35 | 139 | 39 | 13 | 86 | 59 | 24 | 415 | 22 | 111 | 21 | 156 | 95 | 336 | 12 | 36 | 40 |
| **PP37** | 73 | 84 | 22 | 15 | 55 | 155 | 18 | 40 | 28 | 93 | 48 | 3256 | 168 | 566 | 87 | 579 | 44 |
| **PP38** | 17 | 47 | 36 | 18 | 33 | 82 | 48 | 36 | 130 | 143 | 21 | 42 | 236 | 40 | 52 | 119 | 318 |
| **PP39** | 24 | 332 | 11 | 6 | 57 | 54 | 3 | 758 | 19 | 397 | 23 | 37 | 223 | 220 | 17 | 51 | 56 |
| **PP40** | 10 | 134 | 31 | 12 | 60 | 41 | 3 | 12 | 621 | 915 | 10 | 90 | 215 | 25 | 18 | 205 | 68 |
| **PP41** | 12 | 336 | 20 | 7 | 29 | 194 | 225 | 85 | 7018 | 83 | 7 | 39 | 186 | 85 | 182 | 4881 | 118 |
| **PP42** | 57 | 625 | 483 | 303 | 1371 | 41 | 6 | 154 | 50 | 53 | 6 | 412 | 911 | 96 | 33 | 92 | 69 |
| **PP43** | 9 | 351 | 45 | 23 | 173 | 52 | 4 | 30 | 202 | 7094 | 12 | 41 | 883 | 1088 | 172 | 115 | 102 |
| **PP44** | 39 | 1236 | 826 | 469 | 1808 | 282 | 19 | 129 | 142 | 221 | 20 | 145 | 383 | 25 | 25 | 142 | 132 |
| **PP45** | 18 | 100 | 19 | 10 | 52 | 55 | 4 | 20 | 87 | 50 | 9 | 210 | 87 | 293 | 9 | 83 | 97 |
| **PP46** | 24 | 246 | 112 | 61 | 330 | 128 | 5 | 36 | 96 | 216 | 14 | 182 | 3495 | 398 | 17 | 156 | 1132 |
| **PP47** | 94 | 242 | 23 | 5 | 37 | 727 | 8 | 142 | 46 | 257 | 30 | 302 | 71 | 303 | 58 | 1152 | 134 |
| **PP48** | 23 | 940 | 77 | 8 | 38 | 54 | 5 | 88 | 179 | 393 | 44 | 60 | 739 | 377 | 18 | 191 | 298 |
| **PP49** | 21 | 2557 | 20 | 8 | 29 | 151 | 14 | 571 | 275 | 180 | 70 | 305 | 6098 | 115 | 14 | 107 | 260 |
| **PP50** | 45 | 80 | 35 | 15 | 358 | 98 | 6 | 23 | 131 | 735 | 12 | 156 | 46 | 43 | 21 | 262 | 123 |
| **PP51** | 76 | 498 | 32 | 18 | 283 | 204 | 13 | 106 | 200 | 559 | 76 | 2193 | 125 | 471 | 36 | 953 | 276 |
| **PP52** | 2962 | 328 | 663 | 111 | 664 | 124 | 11 | 1919 | 28 | 89 | 83 | 480 | 792 | 1666 | 68 | 69 | 47 |
| ***Systemic lupus erythematosus*** | | | | | | | | | | | | | | | | | |
| **SLE01** | 71 | 609 | 263 | 309 | 1154 | 169 | 9 | 46 | 134 | 293 | 8 | 231 | 367 | 55 | 24 | 69 | 490 |
| **SLE02** | 27199 | 10533 | 106 | 42 | 259 | 154 | 37 | 171 | 563 | 20933 | 525 | 667 | 999 | 14812 | 414 | 736 | 900 |
| **SLE03** | 4441 | 1006 | 486 | 8692 | 23034 | 170 | 48 | 131 | 10008 | 685 | 69 | 283 | 126 | 7248 | 365 | 3078 | 194 |
| **SLE04** | 23398 | 4840 | 40 | 76 | 506 | 88 | 68 | 69 | 144 | 281 | 18 | 111 | 1145 | 683 | 31 | 698 | 142 |
| **SLE05** | 4663 | 19781 | 10928 | 21 | 145 | 132 | 17 | 30 | 274 | 293 | 17 | 58 | 200 | 13378 | 1139 | 1826 | 208 |
| **SLE06** | 603 | 656 | 211 | 42 | 357 | 1040 | 27 | 771 | 16976 | 2255 | 1220 | 729 | 2469 | 2625 | 1208 | 2291 | 680 |
| **SLE07** | 80 | 1447 | 1204 | 778 | 2584 | 182 | 20 | 1273 | 43 | 321 | 46 | 247 | 1280 | 1269 | 47 | 301 | 152 |
| **SLE08** | 23584 | 1120 | 8501 | 458 | 4717 | 181 | 19 | 108 | 191 | 617 | 98 | 1223 | 238 | 18447 | 191 | 1233 | 524 |
| **SLE09** | 36 | 1539 | 132 | 89 | 742 | 160 | 5416 | 114 | 332 | 2846 | 72 | 298 | 1412 | 152 | 674 | 216 | 179 |
| **SLE10** | 62 | 1876 | 1647 | 1122 | 3379 | 61 | 34 | 134 | 85 | 405 | 17 | 4265 | 882 | 476 | 1541 | 549 | 7342 |
| **SLE11** | 45 | 367 | 406 | 345 | 4737 | 243 | 23 | 717 | 556 | 1107 | 27 | 1624 | 489 | 5861 | 167 | 1838 | 1244 |
| **SLE12** | 27020 | 16535 | 8636 | 43 | 435 | 186 | 22 | 96 | 82 | 3839 | 576 | 410 | 1288 | 329 | 449 | 464 | 203 |
| **SLE13** | 868 | 2567 | 3491 | 26763 | 38376 | 2964 | 148 | 2077 | 2532 | 34846 | 2142 | 1529 | 2906 | 17227 | 2184 | 19560 | 6626 |
| **SLE14** | 75 | 1669 | 404 | 529 | 2221 | 586 | 237 | 1273 | 137 | 10195 | 35 | 678 | 433 | 1400 | 155 | 1345 | 3383 |
| **SLE15** | 6411 | 2347 | 1074 | 15992 | 25368 | 325 | 43 | 159 | 522 | 882 | 115 | 707 | 2210 | 12760 | 416 | 444 | 367 |
| **SLE16** | 14549 | 1965 | 7896 | 57 | 1795 | 2654 | 17 | 4383 | 175 | 417 | 30 | 3051 | 1784 | 2119 | 227 | 398 | 530 |
| **SLE17** | 907 | 710 | 344 | 12932 | 30319 | 726 | 30 | 199 | 2299 | 4393 | 42 | 497 | 938 | 6408 | 556 | 8519 | 7069 |
| **SLE18** | 174 | 64 | 29 | 14 | 64 | 129 | 11 | 15 | 17 | 1102 | 30 | 42 | 130 | 134 | 197 | 232 | 58 |
| **SLE19** | 2842 | 999 | 526 | 175 | 2779 | 371 | 21 | 308 | 120 | 46054 | 37872 | 798 | 794 | 8000 | 395 | 5722 | 444 |
| ***Systemic sclerosis*** | | | | | | | | | | | | | | | | | |
| **SSc01** | 39 | 96 | 110 | 17 | 70 | 233 | 12 | 124 | 176 | 317 | 651 | 33353 | 87 | 10542 | 89 | 290 | 513 |
| **SSc02** | 634 | 728 | 192 | 4176 | 1015 | 330 | 21 | 103 | 356 | 543 | 454 | 2172 | 3689 | 786 | 5802 | 416 | 727 |
| **SSc03** | 49 | 1597 | 546 | 877 | 2697 | 137 | 15 | 96 | 53 | 331 | 196 | 2342 | 1030 | 392 | 95 | 423 | 2781 |
| **SSc04** | 6321 | 8726 | 384 | 171 | 1855 | 429 | 26 | 141 | 1202 | 322 | 93 | 718 | 306 | 494 | 1106 | 289 | 667 |
| **SSc05** | 1626 | 185 | 69 | 49 | 347 | 424 | 25 | 93 | 106 | 193 | 70 | 26583 | 177 | 241 | 44 | 108 | 69 |
| **SSc06** | 91 | 10086 | 47 | 1573 | 126 | 193 | 10 | 29 | 27 | 339 | 32 | 2655 | 220 | 93 | 14 | 108 | 150 |
| **SSc07** | 46 | 140 | 83 | 96 | 611 | 112 | 17 | 122 | 183 | 644 | 1482 | 886 | 303 | 1619 | 29 | 493 | 130 |
| **SSc08** | 29800 | 14128 | 27193 | 31 | 292 | 112 | 17 | 130 | 527 | 309 | 64 | 759 | 168 | 263 | 36 | 2761 | 307 |
| **SSc09** | 107 | 192 | 163 | 67 | 452 | 131 | 28 | 81 | 2888 | 975 | 202 | 29348 | 443 | 434 | 435 | 569 | 937 |
| **SSc10** | 60 | 2335 | 66 | 19 | 211 | 64 | 8 | 116 | 12845 | 31602 | 36244 | 281 | 768 | 79 | 29 | 885 | 126 |
| **SSc11** | 47 | 243 | 67 | 15 | 161 | 48 | 102 | 540 | 58 | 97 | 10 | 203 | 4806 | 37 | 18 | 372 | 138 |
| **SSc12** | 34 | 3309 | 56 | 23 | 113 | 142 | 6 | 271 | 87 | 459 | 1269 | 435 | 396 | 3111 | 45 | 204 | 649 |
| **SSc13** | 97 | 1306 | 338 | 412 | 1686 | 161 | 27 | 194 | 393 | 44464 | 38668 | 1378 | 532 | 526 | 81 | 730 | 477 |
| **SSc14** | 272 | 8866 | 80 | 53 | 607 | 362 | 35 | 82 | 68 | 39925 | 37657 | 4214 | 51 | 66 | 97 | 116 | 56 |
| **SSc15** | 37 | 53 | 122 | 16 | 192 | 51 | 30 | 100 | 37 | 249 | 73 | 33493 | 599 | 778 | 63 | 296 | 136 |
| **SSc16** | 92 | 214 | 113 | 47 | 333 | 96 | 4 | 231 | 25 | 326 | 13 | 364 | 92 | 698 | 15 | 81 | 83 |
| **SSc17** | 38 | 67 | 39 | 23 | 213 | 211 | 5 | 52 | 26 | 866 | 91 | 11089 | 141 | 277 | 31 | 108 | 180 |
| **SSc18** | 11234 | 1392 | 362 | 41 | 118 | 134 | 21 | 450 | 235 | 1322 | 74 | 423 | 130 | 153 | 67 | 153 | 1311 |
| **SSc19** | 64 | 182 | 211 | 27 | 220 | 156 | 14 | 71 | 3649 | 1920 | 497 | 554 | 657 | 462 | 47 | 1672 | 3822 |
| **SSc20** | 30 | 176 | 88 | 59 | 243 | 295 | 24 | 81 | 188 | 40572 | 38591 | 250 | 6132 | 887 | 619 | 84 | 84 |
| **SSc21** | 78 | 12979 | 102 | 10393 | 9101 | 157 | 8867 | 112 | 997 | 458 | 98 | 666 | 97 | 435 | 136 | 757 | 533 |
| **SSc22** | 76 | 13229 | 160 | 133 | 174 | 328 | 15 | 493 | 33 | 34908 | 33445 | 158 | 58 | 224 | 39 | 301 | 297 |
| **SSc23** | 15842 | 4346 | 16685 | 63 | 604 | 285 | 18 | 167 | 311 | 294 | 103 | 307 | 817 | 6548 | 53 | 930 | 329 |
| **SSc24** | 33 | 117 | 66 | 13045 | 7711 | 490 | 12 | 104 | 901 | 232 | 51 | 151 | 199 | 71 | 1465 | 237 | 92 |
| **SSc25** | 77 | 199 | 304 | 75 | 197 | 266 | 2329 | 111 | 6961 | 31310 | 34978 | 447 | 313 | 89 | 45 | 687 | 408 |
| **SSc26** | 21 | 81 | 111 | 14 | 176 | 201 | 4 | 189 | 375 | 2201 | 1271 | 201 | 85 | 106 | 42 | 1038 | 239 |
| **SSc27** | 84 | 8075 | 69 | 34 | 327 | 87 | 20 | 172 | 60 | 389 | 8319 | 882 | 20841 | 434 | 71 | 80 | 88 |
| **SSc28** | 349 | 11396 | 319 | 365 | 2906 | 164 | 21 | 298 | 252 | 448 | 100 | 1964 | 601 | 289 | 163 | 84 | 212 |
| **SSc29** | 22 | 142 | 43 | 17 | 174 | 188 | 11 | 72 | 22 | 4624 | 6 | 30731 | 250 | 72 | 52 | 231 | 179 |
| **SSc30** | 31 | 133 | 176 | 40 | 180 | 390 | 22 | 605 | 154 | 5678 | 32 | 864 | 102 | 200 | 41 | 4621 | 102 |
| **SSc31** | 145 | 436 | 222 | 35 | 246 | 101 | 19 | 125 | 30 | 124 | 25 | 516 | 239 | 150 | 61 | 180 | 81 |
| **SSc32** | 11 | 418 | 22 | 9 | 99 | 87 | 2 | 306 | 90 | 156 | 64 | 131 | 36 | 394 | 23 | 210 | 69 |
| **SSc33** | 34 | 143 | 56 | 25 | 167 | 264 | 7 | 191 | 56 | 2699 | 534 | 29444 | 452 | 185 | 39 | 101 | 334 |
| **SSc34** | 41 | 6626 | 73 | 27 | 447 | 98 | 13 | 568 | 515 | 3885 | 541 | 10439 | 302 | 305 | 78 | 27255 | 3131 |
| **SSc35** | 507 | 13729 | 433 | 171 | 3338 | 1140 | 37 | 148 | 196 | 30832 | 33021 | 1355 | 231 | 222 | 84 | 910 | 328 |
| **SSc36** | 29 | 371 | 480 | 1750 | 778 | 68 | 11 | 29 | 209 | 83 | 195 | 898 | 109 | 54 | 31 | 102 | 184 |
| **SSc37** | 59 | 245 | 169 | 24 | 202 | 98 | 183 | 55 | 158 | 224 | 50 | 16062 | 622 | 1943 | 290 | 513 | 200 |
| **SSc38** | 170 | 1538 | 150 | 45 | 444 | 222 | 22 | 195 | 90 | 29318 | 34690 | 1647 | 2709 | 37 | 118 | 4771 | 236 |
| **SSc39** | 2252 | 533 | 182 | 108 | 493 | 136 | 168 | 147 | 265 | 2512 | 109 | 37226 | 598 | 377 | 145 | 710 | 533 |
| **SSc40** | 84 | 297 | 96 | 25 | 112 | 3544 | 253 | 121 | 190 | 675 | 1820 | 26125 | 730 | 965 | 127 | 250 | 440 |
| ***SARS-CoV-2+ INPT <30d PSO*** | | | | | | | | | | | | | | | | | |
| **IP01** | 6897 | 953 | 60 | 69 | 349 | 167 | 37 | 112 | 1015 | 953 | 3371 | 383 | 7097 | 326 | 93 | 513 | 319 |
| **IP02** | 30 | 201 | 102 | 49 | 282 | 221 | 15 | 96 | 94 | 438 | 93 | 206 | 4794 | 1078 | 86 | 1165 | 84 |
| **IP03** | 31 | 276 | 1755 | 23 | 151 | 193 | 27 | 134 | 570 | 476 | 762 | 355 | 2466 | 265 | 53 | 397 | 181 |
| **IP05** | 83 | 146 | 51 | 9 | 54 | 547 | 17 | 90 | 21 | 429 | 226 | 89 | 111 | 300 | 786 | 1091 | 70 |
| **IP06** | 37 | 593 | 47 | 23 | 151 | 138 | 70 | 144 | 3903 | 1194 | 1384 | 961 | 11698 | 643 | 71 | 9164 | 2403 |
| **IP07** | 35 | 114 | 28 | 12 | 53 | 68 | 16 | 31 | 43 | 259 | 33 | 76 | 388 | 293 | 22 | 7503 | 60 |
| **IP08** | 25 | 429 | 29 | 13 | 90 | 44 | 34 | 132 | 89 | 10897 | 31 | 429 | 439 | 137 | 34 | 460 | 168 |
| **IP11** | 17 | 111 | 49 | 11 | 50 | 121 | 18 | 318 | 24 | 217 | 485 | 341 | 79 | 44 | 27 | 126 | 72 |
| **IP13** | 29 | 204 | 42 | 24 | 141 | 206 | 33 | 1200 | 1056 | 6381 | 403 | 2598 | 7249 | 7080 | 80 | 3115 | 157 |
| **IP14** | 32 | 473 | 105 | 28 | 179 | 567 | 101 | 514 | 1547 | 2483 | 152 | 98 | 509 | 202 | 37 | 232 | 305 |
| **IP15** | 37 | 1107 | 56 | 40 | 106 | 469 | 42 | 72 | 66 | 294 | 19 | 124 | 462 | 153 | 45 | 4398 | 309 |
| **IP16** | 26 | 929 | 135 | 51 | 227 | 102 | 15 | 51 | 102 | 1530 | 13 | 1073 | 3668 | 331 | 68 | 1265 | 589 |
| **IP17** | 44 | 21069 | 93 | 314 | 11184 | 118 | 242 | 134 | 534 | 519 | 82 | 763 | 2386 | 3131 | 207 | 134 | 4330 |
| **IP18** | 51 | 982 | 339 | 241 | 2198 | 896 | 31 | 201 | 60 | 3977 | 42 | 292 | 1535 | 1111 | 111 | 10027 | 2743 |
| **IP19** | 206 | 310 | 424 | 39 | 290 | 71 | 14 | 37 | 39 | 1155 | 902 | 469 | 1837 | 313 | 129 | 2185 | 624 |
| **IP20** | 5678 | 2279 | 1736 | 1119 | 4392 | 709 | 50 | 143 | 74 | 835 | 59 | 799 | 1479 | 180 | 191 | 2421 | 4605 |
| **IP21** | 128 | 844 | 129 | 103 | 555 | 397 | 31 | 130 | 87 | 334 | 56 | 618 | 433 | 135 | 105 | 764 | 494 |
| **IP22** | 82 | 2993 | 160 | 209 | 705 | 166 | 58 | 69 | 84 | 240 | 35 | 3058 | 1368 | 4556 | 52 | 281 | 847 |
| **IP23** | 24 | 1634 | 135 | 75 | 465 | 208 | 20 | 106 | 101 | 4590 | 42 | 515 | 654 | 705 | 39 | 740 | 169 |
| **IP24** | 42 | 237 | 29 | 21 | 156 | 107 | 16 | 54 | 98 | 311 | 1511 | 388 | 83 | 106 | 39 | 1532 | 457 |
| **IP25** | 115 | 1273 | 100 | 37 | 243 | 424 | 16 | 59 | 113 | 537 | 46 | 956 | 2937 | 47 | 1201 | 3724 | 628 |
| **IP26** | 34 | 487 | 34 | 34 | 165 | 146 | 16 | 530 | 177 | 2543 | 41 | 139 | 97 | 604 | 120 | 176 | 300 |
| **IP27** | 22 | 75 | 26 | 13 | 90 | 37 | 5 | 27 | 101 | 63 | 734 | 120 | 79 | 27 | 22 | 52 | 46 |
| **IP28** | 18 | 207 | 865 | 16 | 205 | 67 | 8 | 158 | 25 | 10095 | 79 | 1385 | 229 | 29 | 19 | 119 | 59 |
| **IP29** | 87 | 1620 | 475 | 299 | 1286 | 468 | 34 | 223 | 129 | 2055 | 162 | 2099 | 2282 | 15027 | 157 | 912 | 394 |
| **IP30** | 11 | 30 | 8 | 7 | 24 | 25 | 2 | 321 | 6 | 86 | 5 | 108 | 28 | 11 | 11 | 50 | 52 |
| **IP31** | 13 | 58 | 20 | 11 | 37 | 42 | 5 | 81 | 16 | 936 | 6 | 14 | 47 | 24 | 16 | 134 | 73 |
| **IP32** | 83 | 11798 | 763 | 68 | 171 | 182 | 42 | 14590 | 1064 | 795 | 4720 | 1915 | 133 | 640 | 369 | 3131 | 747 |
| **IP33** | 126 | 403 | 286 | 128 | 163 | 759 | 37 | 384 | 170 | 1036 | 30 | 594 | 12111 | 321 | 891 | 1407 | 3004 |
| **IP34** | 28 | 108 | 60 | 22 | 79 | 115 | 10 | 58 | 1037 | 204 | 33 | 1247 | 246 | 114 | 144 | 92 | 821 |
| **IP35** | 30 | 245 | 308 | 70 | 370 | 88 | 18 | 180 | 304 | 1616 | 136 | 680 | 215 | 338 | 118 | 162 | 10010 |
| **IP37** | 52 | 340 | 80 | 52 | 91 | 151 | 207 | 466 | 220 | 521 | 786 | 287 | 328 | 119 | 91 | 640 | 1252 |
| **IP39** | 19 | 369 | 46 | 28 | 124 | 265 | 20 | 140 | 56 | 232 | 22 | 98 | 221 | 436 | 90 | 1145 | 130 |
| **IP40** | 10 | 75 | 14 | 11 | 21 | 567 | 3 | 114 | 23 | 61 | 3 | 98 | 103 | 211 | 48 | 63 | 34 |
| ***SARS-CoV-2+ OUTPT <30d PSO*** | | | | | | | | | | | | | | | | | |
| **OP02** | 92 | 276 | 100 | 63 | 265 | 149 | 72 | 63 | 514 | 2545 | 1604 | 157 | 169 | 2153 | 55 | 311 | 100 |
| **OP06** | 20 | 175 | 189 | 16 | 91 | 70 | 11 | 133 | 74 | 128 | 284 | 140 | 509 | 425 | 164 | 531 | 81 |
| **OP09** | 383 | 313 | 649 | 74 | 873 | 253 | 96 | 185 | 204 | 367 | 785 | 606 | 1019 | 762 | 68 | 340 | 309 |
| **OP15** | 28 | 741 | 22 | 10 | 99 | 269 | 12 | 149 | 80 | 228 | 161 | 187 | 709 | 838 | 44 | 352 | 173 |
| **OP16** | 21 | 533 | 298 | 184 | 895 | 67 | 270 | 58 | 173 | 478 | 26 | 950 | 2096 | 304 | 32 | 573 | 5460 |
| **OP17** | 91 | 483 | 155 | 41 | 213 | 412 | 31 | 74 | 376 | 342 | 5577 | 1042 | 9334 | 1684 | 410 | 304 | 1315 |
| **OP18** | 38 | 142 | 63 | 22 | 211 | 134 | 19 | 157 | 282 | 1026 | 13 | 357 | 530 | 716 | 333 | 145 | 478 |
| **OP19** | 100 | 359 | 109 | 72 | 848 | 127 | 17 | 22 | 51 | 98 | 7 | 318 | 163 | 4141 | 39 | 932 | 131 |
| **OP20** | 46 | 185 | 51 | 18 | 150 | 424 | 87 | 188 | 109 | 204 | 117 | 277 | 168 | 209 | 740 | 517 | 2379 |
| **OP21** | 30 | 260 | 88 | 16 | 135 | 122 | 11 | 70 | 1235 | 319 | 447 | 238 | 1266 | 657 | 97 | 225 | 353 |
| **OP22** | 107 | 283 | 651 | 26 | 235 | 126 | 20 | 325 | 1198 | 1494 | 392 | 961 | 2136 | 5218 | 824 | 337 | 470 |
| **OP23** | 46 | 187 | 51 | 30 | 237 | 259 | 14 | 90 | 91 | 378 | 40 | 130 | 218 | 19094 | 98 | 1619 | 170 |
| **OP25** | 45 | 2233 | 1169 | 757 | 2616 | 144 | 24 | 67 | 215 | 429 | 72 | 1243 | 1293 | 2134 | 139 | 2411 | 235 |
| **OP26** | 39 | 495 | 668 | 182 | 929 | 650 | 14 | 71 | 67 | 412 | 48 | 580 | 2512 | 488 | 33 | 1412 | 250 |
| **OP28** | 57 | 298 | 161 | 71 | 439 | 443 | 30 | 131 | 74 | 162 | 21 | 154 | 237 | 697 | 42 | 187 | 165 |
| **OP29** | 10 | 103 | 5 | 3 | 19 | 73 | 10 | 21 | 64 | 94 | 2 | 92 | 451 | 867 | 17 | 220 | 21 |
| **OP30** | 21 | 368 | 115 | 57 | 445 | 236 | 11 | 49 | 945 | 935 | 38 | 342 | 394 | 117 | 100 | 1235 | 228 |
| **OP33** | 4 | 27 | 5 | 4 | 24 | 47 | 4 | 69 | 27 | 59 | 3 | 34 | 334 | 15 | 7 | 35 | 31 |
| ***SARS-CoV-2+ INPT >90d PSO*** | | | | | | | | | | | | | | | | | |
| **IP02** | 25 | 438 | 1854 | 130 | 715 | 164 | 38 | 141 | 381 | 259 | 15 | 249 | 2351 | 222 | 29 | 230 | 91 |
| **IP04** | 36 | 1154 | 69 | 33 | 229 | 82 | 15 | 317 | 722 | 520 | 14 | 498 | 103 | 532 | 106 | 235 | 978 |
| **IP05** | 196 | 287 | 123 | 11 | 99 | 1154 | 28 | 444 | 39 | 718 | 578 | 150 | 132 | 541 | 1168 | 1900 | 126 |
| **IP06** | 32 | 550 | 32 | 19 | 126 | 133 | 58 | 158 | 577 | 1048 | 329 | 786 | 1016 | 551 | 54 | 5292 | 853 |
| **IP09** | 54 | 117 | 40 | 19 | 153 | 140 | 17 | 29 | 183 | 405 | 1165 | 499 | 366 | 440 | 35 | 628 | 421 |
| **IP10** | 67 | 962 | 252 | 140 | 918 | 84 | 24 | 190 | 762 | 422 | 51 | 624 | 1661 | 170 | 36 | 554 | 1085 |
| **IP12** | 27 | 72 | 67 | 24 | 127 | 82 | 16 | 97 | 31 | 95 | 450 | 79 | 76 | 74 | 17 | 248 | 898 |
| **IP16** | 48 | 978 | 90 | 57 | 379 | 270 | 28 | 86 | 393 | 2080 | 21 | 1486 | 2464 | 8714 | 113 | 1968 | 486 |
| **IP25** | 123 | 2636 | 116 | 63 | 159 | 412 | 27 | 95 | 231 | 947 | 84 | 942 | 4116 | 161 | 1151 | 4576 | 804 |
| **IP28** | 20 | 252 | 892 | 23 | 213 | 80 | 11 | 212 | 28 | 10100 | 124 | 1416 | 255 | 53 | 24 | 144 | 91 |
| **IP29** | 85 | 1448 | 508 | 304 | 1307 | 372 | 34 | 200 | 162 | 1809 | 126 | 2082 | 2235 | 13466 | 152 | 1058 | 2952 |
| **IP30** | 11 | 28 | 7 | 6 | 27 | 17 | 1 | 342 | 5 | 267 | 8 | 161 | 15 | 6 | 8 | 59 | 30 |
| **IP31** | 20 | 53 | 23 | 13 | 51 | 47 | 4 | 54 | 11 | 762 | 12 | 28 | 317 | 21 | 16 | 168 | 144 |
| **IP32** | 119 | 13379 | 989 | 65 | 181 | 171 | 33 | 20250 | 1475 | 838 | 5484 | 2252 | 388 | 627 | 453 | 2753 | 795 |
| **IP33** | 76 | 337 | 137 | 72 | 106 | 625 | 45 | 113 | 299 | 1246 | 30 | 197 | 4261 | 667 | 377 | 1409 | 2180 |
| **IP34** | 28 | 111 | 56 | 25 | 66 | 103 | 13 | 59 | 896 | 189 | 1101 | 1275 | 177 | 57 | 121 | 106 | 560 |
| **IP35** | 38 | 501 | 333 | 195 | 924 | 131 | 25 | 111 | 1304 | 1309 | 64 | 737 | 292 | 483 | 33 | 237 | 3632 |
| **IP36** | 254 | 4441 | 1898 | 30 | 90 | 89 | 16 | 306 | 6166 | 862 | 90 | 71 | 13922 | 158 | 71 | 435 | 24933 |
| **IP39** | 58 | 167 | 177 | 39 | 130 | 247 | 18 | 259 | 51 | 401 | 17 | 318 | 356 | 3550 | 49 | 249 | 674 |
| **IP40** | 5 | 33 | 7 | 5 | 16 | 449 | 2 | 35 | 10 | 55 | 1 | 48 | 143 | 53 | 11 | 12 | 16 |
| ***SARS-CoV-2+ OUTPT >90d PSO*** | | | | | | | | | | | | | | | | | |
| **OP01** | 19 | 81 | 42 | 9 | 17 | 78 | 13 | 148 | 27 | 499 | 12 | 10779 | 43 | 35 | 61 | 1627 | 69 |
| **OP02** | 66 | 221 | 75 | 46 | 197 | 116 | 49 | 51 | 353 | 1878 | 1201 | 111 | 77 | 1888 | 34 | 93 | 66 |
| **OP03** | 77 | 452 | 103 | 36 | 77 | 201 | 24 | 58 | 63 | 2110 | 101 | 219 | 213 | 184 | 30 | 647 | 225 |
| **OP04** | 71 | 117 | 60 | 22 | 101 | 136 | 27 | 40 | 59 | 3283 | 6 | 1015 | 62 | 339 | 59 | 192 | 106 |
| **OP05** | 24 | 894 | 564 | 343 | 31 | 150 | 15 | 80 | 56 | 1193 | 48 | 144 | 449 | 55 | 37 | 1596 | 112 |
| **OP06** | 19 | 94 | 47 | 15 | 34 | 69 | 9 | 114 | 67 | 105 | 184 | 154 | 299 | 380 | 156 | 540 | 71 |
| **OP07** | 76 | 268 | 85 | 62 | 105 | 649 | 114 | 273 | 48 | 154 | 7 | 2687 | 179 | 124 | 142 | 1548 | 130 |
| **OP08** | 11 | 448 | 19 | 5 | 12 | 117 | 8 | 60 | 217 | 113 | 5 | 346 | 582 | 58 | 103 | 350 | 323 |
| **OP09** | 469 | 324 | 667 | 74 | 193 | 292 | 101 | 206 | 218 | 373 | 286 | 1407 | 1215 | 976 | 70 | 445 | 324 |
| **OP10** | 24 | 630 | 401 | 239 | 34 | 565 | 15 | 72 | 203 | 139 | 149 | 176 | 414 | 3744 | 97 | 809 | 56 |
| **OP11** | 35 | 62 | 20 | 11 | 27 | 304 | 8 | 84 | 260 | 102 | 93 | 3729 | 82 | 209 | 396 | 76 | 82 |
| **OP12** | 47 | 253 | 155 | 83 | 11 | 63 | 9 | 86 | 478 | 894 | 7 | 92 | 99 | 1149 | 6 | 49 | 123 |
| **OP13** | 55 | 3238 | 92 | 31 | 130 | 123 | 17 | 200 | 626 | 3477 | 123 | 844 | 104 | 569 | 28 | 1560 | 152 |
| **OP14** | 44 | 253 | 100 | 53 | 33 | 131 | 16 | 92 | 1647 | 528 | 12 | 1235 | 3912 | 243 | 31 | 735 | 65 |
| **OP15** | 22 | 674 | 16 | 9 | 32 | 236 | 11 | 93 | 67 | 203 | 135 | 104 | 625 | 811 | 39 | 280 | 164 |
| **OP16** | 21 | 495 | 299 | 176 | 43 | 69 | 214 | 53 | 125 | 338 | 26 | 761 | 1839 | 259 | 38 | 477 | 5334 |
| **OP17** | 79 | 438 | 164 | 36 | 43 | 422 | 25 | 82 | 326 | 278 | 5596 | 1005 | 9133 | 1589 | 322 | 247 | 1192 |
| **OP18** | 44 | 173 | 83 | 27 | 58 | 148 | 12 | 177 | 85 | 1199 | 19 | 423 | 663 | 847 | 418 | 181 | 561 |
| **OP19** | 108 | 355 | 105 | 72 | 141 | 137 | 16 | 20 | 53 | 103 | 7 | 376 | 168 | 4153 | 38 | 900 | 163 |
| **OP21** | 41 | 273 | 93 | 19 | 60 | 134 | 37 | 112 | 4889 | 378 | 404 | 367 | 1975 | 883 | 126 | 337 | 450 |
| **OP22** | 84 | 219 | 471 | 19 | 296 | 96 | 16 | 275 | 919 | 1180 | 259 | 754 | 1594 | 2512 | 625 | 252 | 361 |
| **OP23** | 50 | 187 | 66 | 35 | 101 | 501 | 13 | 101 | 127 | 369 | 42 | 131 | 267 | 18103 | 99 | 1720 | 157 |
| **OP24** | 48 | 1138 | 59 | 15 | 89 | 76 | 16 | 58 | 1175 | 914 | 28 | 149 | 265 | 1319 | 1854 | 827 | 1067 |
| **OP25** | 53 | 2467 | 1233 | 832 | 249 | 146 | 22 | 62 | 283 | 445 | 75 | 1106 | 1071 | 2301 | 128 | 1256 | 221 |
| **OP26** | 33 | 422 | 503 | 139 | 122 | 562 | 10 | 46 | 37 | 165 | 31 | 641 | 2075 | 291 | 29 | 1320 | 217 |
| **OP27** | 58 | 350 | 316 | 45 | 431 | 126 | 25 | 72 | 103 | 547 | 855 | 586 | 180 | 556 | 145 | 781 | 348 |
| **OP28** | 57 | 744 | 510 | 298 | 320 | 363 | 25 | 121 | 69 | 144 | 20 | 265 | 374 | 540 | 43 | 150 | 168 |
| **OP29** | 30 | 270 | 26 | 8 | 25 | 176 | 20 | 58 | 179 | 340 | 11 | 291 | 1418 | 1823 | 44 | 637 | 65 |
| **OP30** | 14 | 214 | 67 | 37 | 28 | 165 | 10 | 35 | 651 | 645 | 24 | 260 | 273 | 69 | 63 | 549 | 138 |
| **OP31** | 22 | 250 | 44 | 10 | 30 | 104 | 8 | 259 | 72 | 291 | 2066 | 303 | 217 | 1299 | 49 | 913 | 170 |
| **OP32** | 80 | 171 | 68 | 29 | 356 | 577 | 27 | 168 | 1530 | 1885 | 11 | 231 | 116 | 245 | 526 | 346 | 503 |
| **OP33** | 17 | 102 | 18 | 12 | 22 | 219 | 14 | 206 | 350 | 185 | 35 | 159 | 1855 | 98 | 41 | 247 | 173 |

Supplemental Figure 3

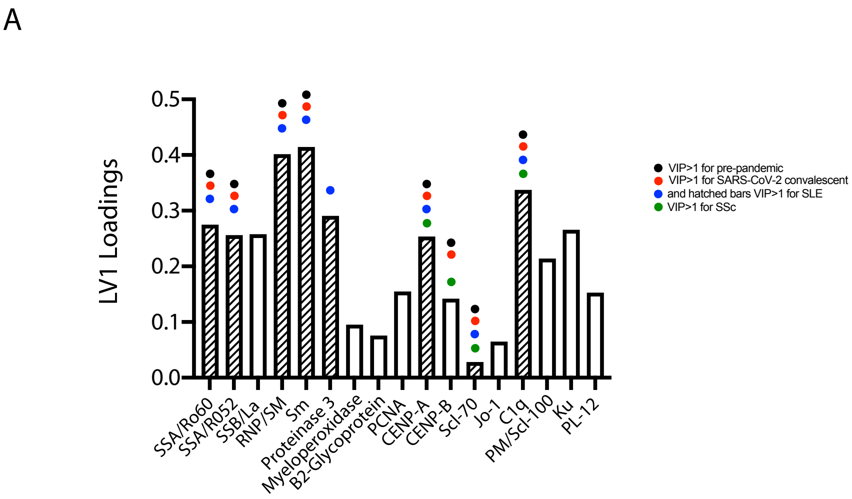


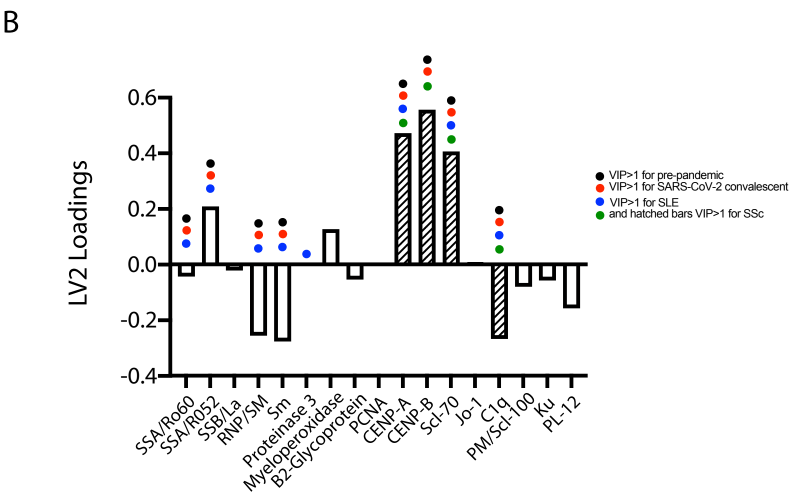

**Supplemental Figure 4.**

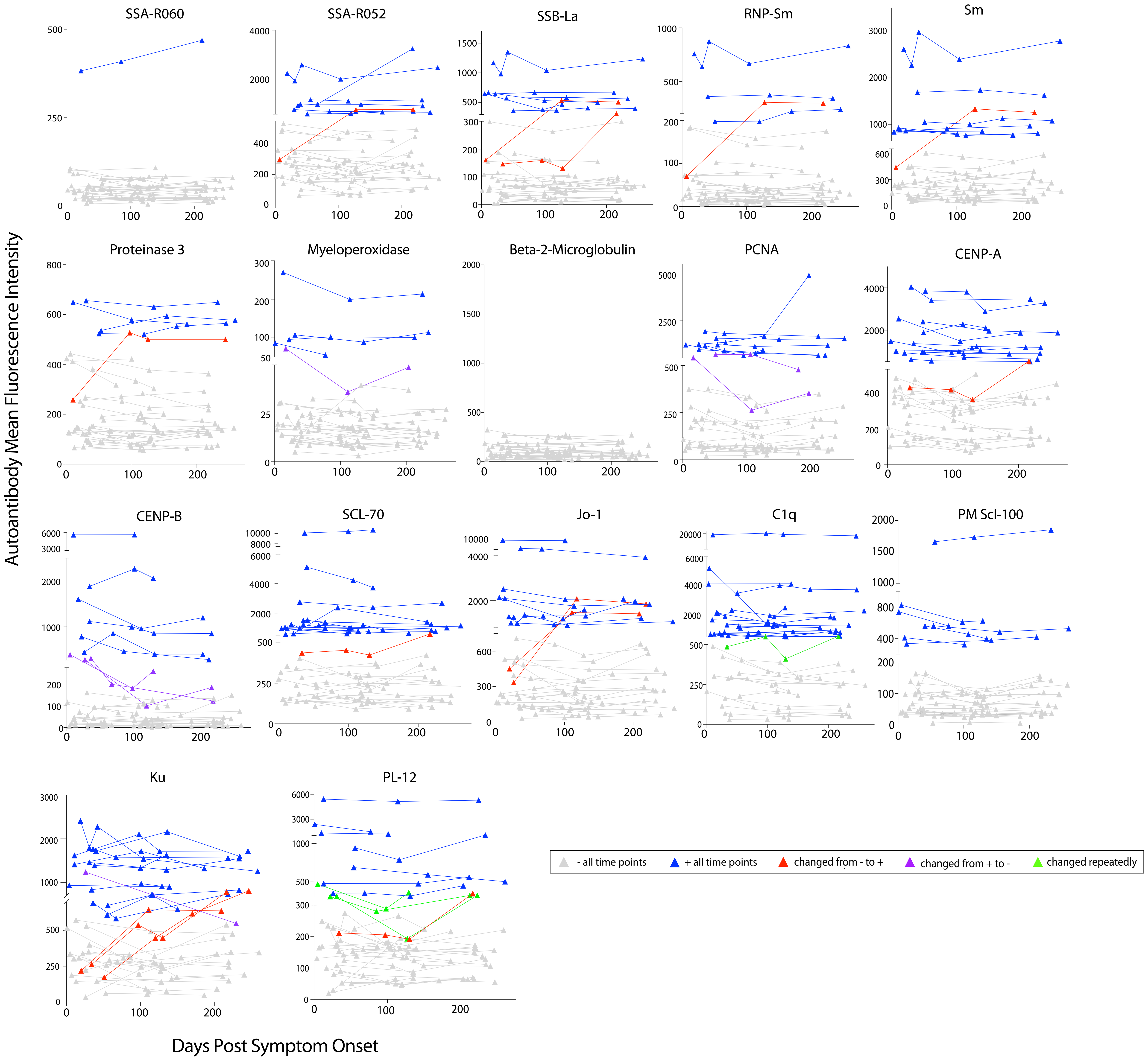

**Supplemental Figure 5.**

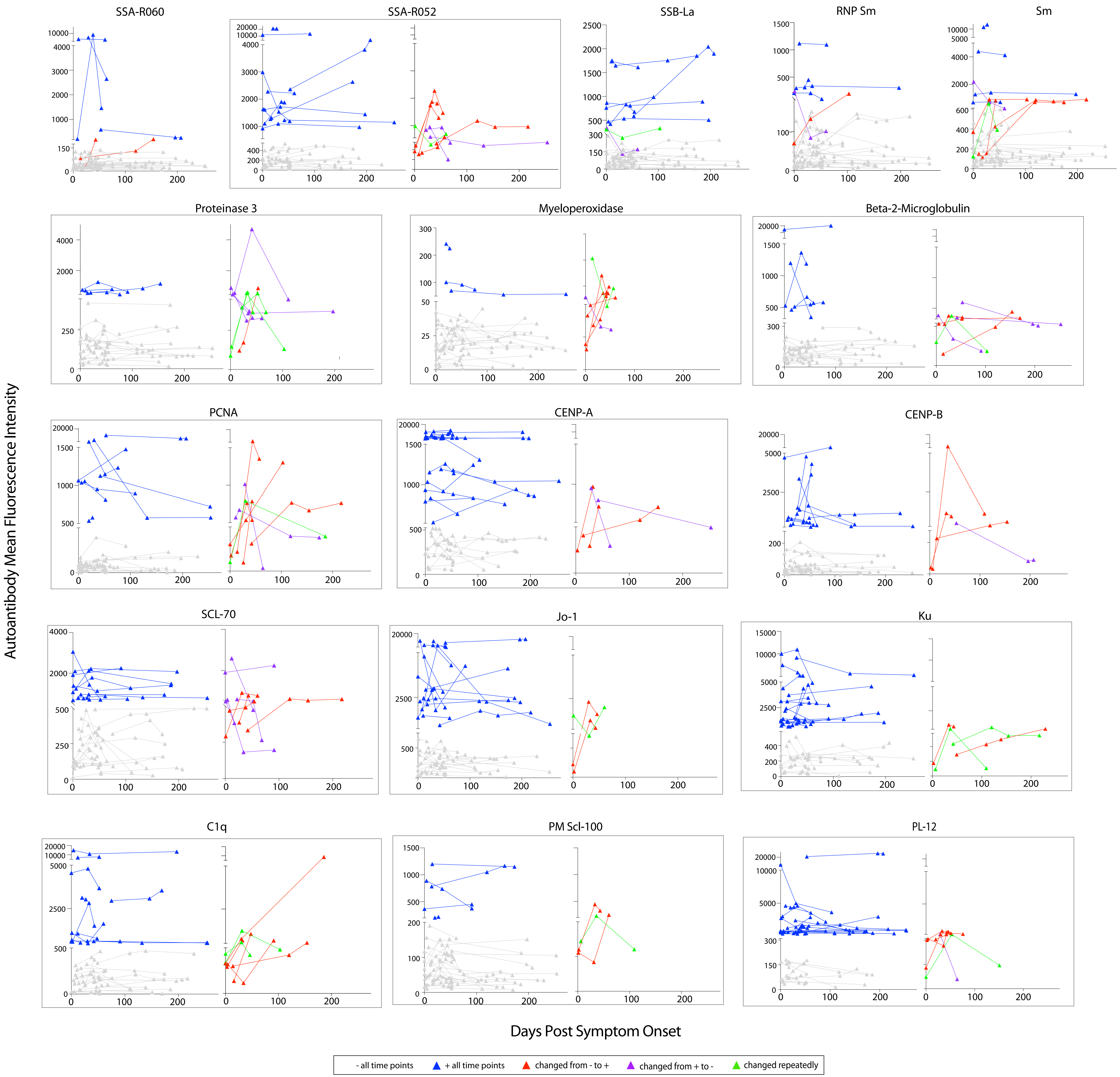
